# Integrative multi-omics characterization of hepatocellular carcinoma in Hispanic patients

**DOI:** 10.1101/2024.04.27.24306447

**Authors:** Debodipta Das, Xiaojing Wang, Yu-Chiao Chiu, Hakim Bouamar, Francis E. Sharkey, Jorge E. Lopera, Zhao Lai, Susan T. Weintraub, Xianlin Han, Yi Zou, Hung-I H. Chen, Carla R. Zeballos Torrez, Xiang Gu, Matyas Cserhati, Joel E. Michalek, Glenn A. Halff, Yidong Chen, Siyuan Zheng, Francisco G. Cigarroa, Lu-Zhe Sun

## Abstract

**Background:** The incidence and mortality rates of hepatocellular carcinoma (HCC) among Hispanics in the United States are much higher than those of non-Hispanic whites. We conducted comprehensive multi-omics analyses to understand molecular alterations in HCC among Hispanic patients.

**Methods:** Paired tumor and adjacent non-tumor samples were collected from 31 Hispanic HCC in South Texas (STX-Hispanic) for genomic, transcriptomic, proteomic, and metabolomic profiling. Additionally, serum lipids were profiled in 40 Hispanic and non-Hispanic patients with or without clinically diagnosed HCC.

**Results:** Exome sequencing revealed high mutation frequencies of *AXIN2* and *CTNNB1* in STX Hispanic HCCs, suggesting a predominant activation of the Wnt/β-catenin pathway. The *TERT* promoter mutation frequency was also remarkably high in the Hispanic cohort. Cell cycles and liver functions were identified as positively- and negatively-enriched, respectively, with gene set enrichment analysis. Gene sets representing specific liver metabolic pathways were associated with dysregulation of corresponding metabolites. Negative enrichment of liver adipogenesis and lipid metabolism corroborated with a significant reduction in most lipids in the serum samples of HCC patients. Two HCC subtypes from our Hispanic cohort were identified and validated with the TCGA liver cancer cohort. The subtype with better overall survival showed higher activity of immune and angiogenesis signatures, and lower activity of liver function-related gene signatures. It also had higher levels of immune checkpoint and immune exhaustion markers.

**Conclusions:** Our study revealed some specific molecular features of Hispanic HCC and potential biomarkers for therapeutic management of HCC and provides a unique resource for studying Hispanic HCC.

## Background

Liver cancer is the sixth most commonly diagnosed cancer, with the third highest mortality rate among all cancers worldwide [1]. For all stages combined, the 5-year survival is the second lowest (∼21%) for liver cancer across all races in the US [2]. Hepatocellular carcinoma (HCC) accounts for the majority (> 90%) of all primary liver cancers [3]. Texas residents have the highest age-adjusted HCC incidence rate, which was estimated to be 45% higher than the US national average. Texas-based Hispanics were the major contributor to this high rate [4]. Though HCC incidence rates started declining very recently in the US, Hispanics had the highest rate among ethnic groups [5]. Earlier studies have implicated liver metabolic disorders associated with obesity and diabetes, which are prevalent in Hispanics, as potential contributors to the high incidence of HCC [6–9]. However, the underlying molecular alterations that contribute to hepatocarcinogenesis remain to be elucidated. Ancestral genetic and epigenetic traits are known to contribute to cancer incidence and prognosis [10], but the factors, including the differences in the genomic, epigenomic, and metabolomic features contributing to the elevated risk of HCC development among Hispanics, are poorly understood.

Recently, there have been large-scale multi-omic studies on HCC to identify the molecular mechanisms for hepatocarcinogenesis. Genomic profiling of HCC identified recurrent somatic mutations in *TP53*, *CTNNB1*, *AXIN1*, and *TERT* promoter [11–14]. These studies have shown frequent mutations and copy number changes in the various driver genes leading to aberrant RTK/RAS/PI3K, TP53/cell cycle, Wnt/β-catenin, AKT/mTOR, Notch pathways, and DNA damage regulation in HCC. Transcriptome, proteome, and phosphoproteome-based molecular subclasses of HCC have shown significant differences in their clinicopathological characteristics [11,13,14]. Classification of HCC based on immune-related gene signatures and pathways provided insights into immunotherapy response and patient prognosis [15,16]. Other groups have studied the metabolomic and lipid profiles of HCC in patient tissue and serum samples [17–19]. While published studies have reported proteogenomic or metabolomic/lipidomic approaches to characterize and cluster HCC tumors of different etiologies and clinical stages in various ethnic groups, integrative analyses of HCC tumors involving both proteogenomic and metabolomic/lipidomic approaches appear lacking.

We performed comprehensive analyses of genomic, transcriptomic, proteomic, and metabolomic profiles of paired tumor and adjacent non-tumor samples of Hispanic HCC patients from South Texas (STX-Hispanics) to investigate specific and common features of Hispanic HCC. Serum lipids were profiled in STX-Hispanic and STX-non-Hispanic patients with or without clinically diagnosed HCC. Our study provides a unique resource of multi-omics data sets from STX-Hispanic HCC patients.

## Methods

### Study population and patient ancestry

The discovery cohort comprised 31 HCC patients of self-reported Hispanic origin recruited from South Texas (STX-Hispanic). The tumor and adjacent non-tumor tissue samples were collected from each patient. A pathologist reviewed each normal sample to ensure no contamination of tumor cells. Due to insufficient samples or poor RNA quality, we were not able to obtain all omics data from every patient sample in our discovery cohort. Additional tumor tissues were collected from an independent set of 38 STX-Hispanic HCC patients (validation cohort).

We confirmed the genetic ancestry of 27 HCC patients (discovery cohort) that underwent WES using a panel of 250 ancestry informative markers (AIMs) carefully designed within exon regions from selected panels of genomic data from 1000 Genome Project [20]. Detailed methods have been described elsewhere [21].

Serum samples were collected from unrelated 40 local patients with (n=20) or without (n=20) clinically diagnosed HCC. Within each group, 10 patients were Hispanic (STX-Hispanic) and the other 10 patients belonged to other ethnicities (STX-Non-Hispanic). Written informed consent was obtained from all participants recruited in this study. The study was approved by UT Health San Antonio Institutional Review Board.

### DNA extraction and whole-exome sequencing (WES)

Genomic DNA was extracted from paired tumor and adjacent non-tumor samples of 27 patients (discovery cohort) and tumor-only samples of unrelated 38 patients (validation cohort) of Hispanic origin (STX-Hispanic). Fresh tumor and non-tumor liver tissues were sliced into small pieces of 2-3 mm^3^ and stored at −80 °C. Genomic DNA was isolated from frozen liver tissues (∼10-20 mg) using QIAamp DNA Mini (Cat #51304) after being mechanically disrupted in a sterile glass homogenizer. The quality of the genomic DNA was confirmed by an agarose gel.

The TruSeq Rapid Exome Library Prep kit (Illumina, CA) was used to capture about 45 Mb of all (∼ 99.5% of NCBI RefSeq) coding regions of the human genome. The 100 bp paired-end sequencing of every 6-plex whole exome library pool was performed on a Illumina HiSeq 3000 system according to the manufacturer’s recommended protocol to achieve an average coverage of ∼100X. The library preparation and sequencing were done at the institutional Genome Sequencing Facility. The paired-end reads were aligned to the human reference genome (hg19) with decoy sequences (as used in the 1000 Genomes Project) using BWA (v0.7.5a) software [22]. The duplicate reads were marked and subsequently removed using Picard (v1.140) (https://broadinstitute.github.io/picard/) and SAMtools (v0.1.19) [23], respectively. Local realignment around insertions/deletions (indels) and base quality recalibration were performed using GATK (v3.6) [24].

### Somatic variant analysis

Two somatic mutation callers, VarScan2 (v2.3.9) [25] and MuTect2 [26], were used to detect somatic variants. Somatic variants detected by VarScan2 were further filtered by variant allele frequency (VAF) ≥ 0.1 and a minimum depth of coverage of 30 in tumor samples. MuTect2 module of GATK was run in tumor-normal mode with default settings to detect somatic variants. All the variants were annotated using ANNOVAR [27]. We considered variants, including SNVs (single-nucleotide variants) and indels, identified by both callers in our analyses. The combined Mutation Annotation Format (MAF) file from all patients was used as an input for the MutSigCV (v4.41) [28] to identify significantly mutated genes.

For tumor-only samples, only varScan2 caller was used (mpileup2cns option) for both SNP and indel calling per tumor sample. The Variant Call Format (VCF) files were combined and annotated using ANNOVAR similar to the somatic variant analysis pipeline.

### Validation of somatic mutations from selected genes using DuplexSeq

For a selected 10 HCC samples with *MTOR* and *AXIN2* mutations detected by WES, the extracted DNA was used for Duplex sequencing library preparation according to TwinStrand’s protocols (TwinStrand Biosciences Inc., Seattle WA, USA). Briefly, 200 ng of DNA was prepared by ultrasonically shearing to a mean fragment size of ∼300 bp followed by end-polishing, A-tailing and ligating to Duplex Sequencing Adapters (Custom designed to cover *MTOR* and *AXIN2* genes, for a total of 27 exon regions, 6,321bp covered footprint). After initial PCR amplification, target regions were enriched using a pool comprising 120-nucleotide biotinylated oligonucleotides following TwinStrands’ instruction.

Prepared libraries were sequenced on the HiSeq 3000 (Illumina, CA) using an average of 89.8 million raw reads per sample. Resulting sequence data were demultiplexed into FASTQ and processed using “TwinStrand Duplex FASTQ to VCF Parallel App” hosted on the DNAnexus platform (v3.17.1). The analysis pipeline includes Duplex Tag extraction, alignment to the UCSC hg38 genome and then grouping the reads by unique molecular identifiers (UMIs, maximum on target coverage >7000 fold), performing duplex consensus calling and post-processing, and finally variant calling as described in [29]. VCF file for each HCC samples were further organized and compared to the corresponding whole exome sequences to confirm the mutations reported for *MTOR* and *AXIN2* genes.

### Comparison of mutation frequencies across the ethnic population

The mutation and clinical data for TCGA samples were obtained from the TCGA Pan-Cancer Atlas (https://gdc.cancer.gov/about-data/publications/pancanatlas). To compare with our discovery cohort, we used non-synonymous somatic mutations of 356 LIHC (liver cancer) patients from the TCGA. Cases without ethnicity information were excluded from the analysis. Additionally, we obtained ICGC liver cancer mutation data from the ICGC PCAWG data portal (https://dcc.icgc.org/) [30]. Non-synonymous SNVs and indels were used for comparing mutation rates of cancer genes across the four cohorts.

### TERT promoter sequencing

The specific primer for TERT promoter region was designed as previously published [31] following Illumina overhang primer strategy to enable direct sequencing of PCR products. Primer sequences are:

`[Forward] TCGTCGGCAGCGTCAGATGTGTATAAGAGACAGCAGCGCTGCCTGAAACTC, and [Reverse] GTCTCGTGGGCTCGGAGATGTGTATAAGAGACAGGTCCTGCCCCTTCACCTT.`

Amplification with overhang primer sequence was done using KAPA HiFi HotStart PCR kit (Roche Diagnostics). Briefly, a 20-µl reaction mixture containing 10 ng genomic DNA and 1 µM final concentration of forward and reverse primers. PCR amplification was carried out according to the following protocol: initial denaturation for 3 min at 95°C, followed by 30 cycles of denaturation at 98°C for 30s, annealing at 57°C for 30s, and elongation at 72°C for 30s, with the final extension step at 72°C for 5 mins. PCR products were examined by Fragment Analyzer and purified by AMPure beads at a ratio of 1.8: 1 (beads to PCR products) and an elution volume of 40 µl. Nextera XT index was added during the second PCR step according to the Illumina protocol. The final PCR products went through one round of AMPure beads purification (1:1) to ensure clean products for sequencing. PCR products were sequenced with 100 bp paired-end sequencing using Illumina HiSeq 3000 sequencer (Illumina, San Diego, CA) with an average of about 1.8M reads per sample. Sequence reads were aligned to the TERT promoter sequence using BWA followed by VarScan2 (v2.3.9) with the following parameters (`mpileup2snp --min-coverage 100 --min-var-freq 0.005 --strand-filter 0`) to detect SNPs with at leave 100-fold coverage and minimal variant frequency at 0.5% in at least one HCC sample (either tumor or adjacent no-tumor sample).

### Identification of mutational signatures

The VCF files containing somatic variants (VAF ≥ 0.1) were read into R/Bioconductor based MutationalPatterns (v3.0.1) [32] package and annotated with the contexts of single nucleotide variants (SNVs). The 96 classes of single base substitution (SBS) mutation signatures were accessed from the publication by Alexandrov et al. [33]. We selected 49 (of 67) SigProfiler SBS mutational signatures, found to be the signatures of potential biological origin [33], as our reference signature matrix. We performed signature decomposition to identify the contributions of 49 signatures for each tumor sample. A signature refit was performed using MutationalPatterns bootstrapping method with 1000 iterations to avoid signature misattribution. The signature decomposition from 1000 replicates was used to calculate the percentage and mean contributions of all signatures with non-zero contributions (where contributions > 0).

### Somatic copy number alteration (SCNA) analysis

We applied the copy number module of the VarScan2 package, with default parameters, to identify the preliminary somatic copy number changes from WES. Adjustment for GC content to filter out minimum region size was performed using the copycaller command of VarScan2, with default parameters. The circular binary segmentation (CBS) algorithm was applied using the R/Bioconductor-based DNAcopy package to find segmented regions. Finally, we applied the GISTIC2.0 (v 2.0.23) algorithm [34] on merged segmented files using the GISTIC_2.0 GenePattern web module (https://cloud.genepattern.org/), with default parameters. The copy number of the DNA segments found amplified/deleted with a q-value cutoff of 0.25 and confidence level ≥ 0.9 were considered as significantly altered SCNAs in STX-Hispanic HCC. A threshold value (GISTIC 2.0) of +/- 2 was considered as high-level copy number gain (amplification)/loss (deletion) for a gene, respectively.

### Identification of frequently altered oncogenic signaling pathways based on mutation data

We downloaded the 10 oncogenic signaling pathways from the study by Sanchez-Vega et al. [35]. All annotated somatic SNVs, small indels, and significantly altered CNAs in our 27 STX-Hispanic HCC cohort were used to filter out putative passenger mutations and CNAs based on the curated list of oncogenic and clinically actionable mutations compiled in the OncoKB database [36]. We have only considered the oncogenic/likely oncogenic/predicted oncogenic somatic alterations. Furthermore, known oncogenes and tumor suppressor genes (OncoKB database) found within the recurrent copy number amplified, and deleted regions, respectively, were retained. As an additional validation step, each gene (with significant CNAs) should have positive correlation between copy-number status and gene expression from HCC patients with both mutation and transcriptome data. The selected genes were mapped to identify all oncogenic pathways altered in STX-Hispanic HCC.

### RNA sequencing and differential gene expression analysis

Total RNA was isolated from the tumor and adjacent non-tumor tissue samples of 13 STX-Hispanic HCC patients using RNA Mini Spin Column of Enzymax LLC (Lexington, KY) according to the manufacturer’s protocol. The quality of RNA samples was assessed using Agilent 2100 Bioanalyzer (Agilent Technologies, CA). Samples with RNA Integrity Number (RIN) ≥ 7.0 was used for the preparation of RNA sequencing libraries using the poly(A)-based TruSeq Stranded mRNA Library Prep kit (Illumina, CA) according to the manufacturer’s protocol. The sequencing libraries were pooled and 100 bp paired-end sequencing was performed on HiSeq 2000, HiSeq 3000, or NovaSeq 6000 Systems (Illumina, CA). The library preparation and sequencing process was done at the institutional Genome Sequencing Facility.

The paired-end reads were aligned to the human reference transcript (build hg19) and reference genome (hg19 with decoy sequence) using TopHat2 (v2.0.8b) [37], with default parameters. The alignment (BAM) files were processed using Expectation Maximization (RSEM) (v1.2.31) algorithm [38] to quantify the expression level of genes as expected counts and normalized expression as FPKM (fragments per kilobase of transcript per million mapped reads). The RSEM counts from paired tumor-nontumor samples were analyzed using the R/Bioconductor-based DESeq2 (v1.30.1) and edgeR (v3.30.3) packages [39,40] to identify the differentially expressed genes. We considered results in agreement with both pipelines. Genes with significant (corrected p-value<0.05) dysregulation by at least 2-fold change (i.e. |log2 FC|L≥L1), in tumor compared to adjacent non-tumor samples, were defined as significantly differentially expressed genes (DEGs). To identify genes potentially associated with HCC tumorigenesis, we have reported DEGs found considerably expressed (average FPKM > 1) in our cohort of 13 STX-Hispanic HCC.

### Fusion gene detection

Fusion events were identified using PRADA2 (v2018) [41], STAR-Fusion (v1.9.0) [42] with default parameters. The Trinity Cancer Transcriptome Analysis Toolkit (CTAT) genome library (Apr062020) was used to run STAR-Fusion. We removed fusion events found in both HCC tumor and adjacent non-tumor tissues. Fusion genes nominated by both algorithms were considered for further analysis.

### Detection of alternative splicing (AS) events

The RNA-seq reads aligned by TopHat2 (v2.0.8b) were used to run rMATS (v4.0.2) [43] for identifying AS events in HCC tumors (excluding HCC_33, HCC_164, and HCC_165). An AS event - skipped exon (SE), mutually exclusive exons (MXE), alternative 5′ splice site (A5SS), alternative 3′ splice site (A3SS), and retained intron (RI) – with FDR corrected p-value<0.05 and |IncLevelDifference|>0.1 were considered significant. We further used the MISO (Mixture of Isoforms) (v0.5.4) framework [44] to identify AS events across all samples. Insert length for each paired-end library was computed before the MISO run by setting other parameters at default. Only those significant AS events also identified by MISO in one or more HCC tumors were validated.

### Curation of TCGA RNA-Seq data

The gene expression quantification (RSEM and FPKM) and clinical data for liver cancer (LIHC) patients from the TCGA study were downloaded using the R/Bioconductor based TCGAbiolinks (v2.18.0) package [45]. The gene expression from paired tumor and adjacent normal tissues were available for 50 (out of 371) TCGA-LIHC patients.

### Sample preparation and mass spectrometry analysis of proteomic data

Approximately 3 mg of tissue (paired tumor and adjacent non-tumor) samples were separately homogenized in a buffer containing 10% SDS/50 mM triethylammonium bicarbonate (TEAB) in the presence of protease and phosphatase inhibitors (HALT; Thermo Scientific) and nuclease (Pierce™ Universal Nuclease for Cell Lysis; Thermo Scientific) in a Barocycler (Pressure BioSciences) for 60 cycles at 35 °C. The homogenates were centrifuged at 21,000 g for 10 min and the supernatants were removed. Aliquots corresponding to 100 µg protein (EZQ™ Protein Quantitation Kit; Thermo Scientific) were reduced with tris(2-carboxyethyl)phosphine hydrochloride (TCEP), alkylated in the dark with iodoacetamide and applied to S-Traps (mini; Protifi) for tryptic digestion (sequencing grade; Promega) in 50 mM TEAB. Peptides were eluted from the S-Traps with 0.2% formic acid in 50% aqueous acetonitrile and quantified using Pierce™ Quantitative Fluorometric Peptide Assay (Thermo Scientific). Data-independent acquisition mass spectrometry (DIA-MS) was conducted on an Orbitrap Fusion Lumos mass spectrometer (Thermo Scientific). On-line HPLC separation was accomplished with an RSLC NANO HPLC system (Thermo Scientific/Dionex: column, PicoFrit™ (New Objective; 75 μm i.d.) packed to 15 cm with C18 adsorbent (Vydac; 218MS 5 μm, 300 Å); mobile phase A, 0.5% acetic acid (HAc)/0.005% trifluoroacetic acid (TFA) in water; mobile phase B, 90% acetonitrile/0.5% HAc/0.005% TFA/9.5% water; gradient 3 to 42% B in 120 min; flow rate, 0.4 μl/min. Peptide aliquots (2-µg) were analyzed using gas-phase fractionation and six 4-m/z windows (30k resolution for precursor and product ion scans, all in the orbitrap) to create an empirically-corrected DIA chromatogram library [46] by searching against a panhuman spectral library comprised of 139,449 proteotypic peptides and 10,316 proteins [47]. Experimental samples were blocked by subject and randomized by tissue designation for sample preparation and analysis; injections of 1.5 or 2 µg of peptides and a 2-h HPLC gradient were employed. MS data for experimental samples were acquired in the orbitrap using 12-m/z windows (staggered; 30k resolution for precursor and product ion scans) and searched against the chromatogram library. Scaffold DIA (v1.3.1; Proteome Software) was used for all DIA-MS data processing. Quartile normalization was applied on the log10 peptide intensities across replicates. Reports of protein identification metrics and normalized intensities were exported directly from Scaffold DIA.

### Identification of differentially abundant proteins

We identified abundances of 6229 proteins from tumor-nontumor paired samples of 15 STX-Hispanic HCC. We performed a paired sample two-tailed t-test using R package (v4.0.4) to compare differential abundances of 4274 proteins (with finite expression at least in 8 of 15 patients) in tumors compared to non-tumor samples. Expression values were log2 transformed before performing the t-test. Proteins with corrected p-value<0.05 and average fold-change ≥ 2 (i.e. average |log2 (fold-change)| ≥ 1) in tumor compared to adjacent non-tumor samples were considered as significantly differentially abundant proteins (DAPs).

### Gene Set Enrichment Analysis (GSEA)

The list of expressed (average FPKM > 1) genes from 13 STX-Hispanic HCC were ranked using average log2 (fold-change) in tumor compared to paired non-tumor samples. The ranked genes were used to run the GSEA algorithm with R package clusterProfiler (v3.18.1) [48] for assessing the enrichment of 50 hallmark gene set collections of the Molecular Signatures Database (MSigDB v7.4) [49]. Enrichment analysis (GSEA) was also performed for tumor-normal paired samples from 50 TCGA-LIHC in the similar method described above. Furthermore, we also applied GSEA algorithm to the ranked list of proteins identified from 15 STX-Hispanic HCC cases. All parameters were set at default while performing GSEA using clusterProfiler.

### Hoshida molecular subtypes of STX-Hispanic HCC

Prediction of the Hoshida’s molecular subtypes [50] was performed for all STX-Hispanic HCC tumors samples using nearest template prediction method with transcriptome or proteome data in R-based CMScaller package (v0.9.2). For proteome data, only detected proteins available in our datasets were considered. Hoshida’s HCC subclass was assigned to each tumor samples with significant (FDR<0.05) confidence.

### Transcriptome and proteome-based classification of HCC

The list of genes used for GSEA analyses described above was ranked using log2 (fold-change) between tumor and paired non-tumor tissues for each STX-Hispanic HCC (n=13) and TCGA-LIHC (n=50). Similarly, we ranked the list of proteins for each HCC patient (STX-Hispanic). Single-sample gene set enrichment analysis (ssGSEA) to investigate the enrichment of 50 hallmark gene sets was performed with the R-based ssGSEA2.0 package (https://github.com/broadinstitute/ssGSEA2.0) using the ranked genes or proteins. Like other studies [15], we used enrichment scores for patient clustering. Unsupervised clustering of STX-Hispanic and TCGA HCC patients was done with Ward’s agglomeration method using correlation distance of the normalized enrichment scores (NES) in R-based pheatmap package (v1.0.12). The 50 hallmark gene sets were similarly clustered based on Euclidean distance calculated from 13 STX-Hispanic HCC.

### Characterization of the immune microenvironment

To analyze the infiltration level of immune cells in each HCC tumor from STX-Hispanics (n=13) and non-Hispanics from TCGA-LIHC (n=50), we applied the ESTIMATE algorithm [51] using the R-based estimate package (v1.0.13). We used the CIBERSORT algorithm [52] to deconvolute the transcriptome profiles of all tumor and non-tumor samples into 22 immune cell-type (LM22) specific scores. A total of 1000 permutations were applied to generate the p-values. ssGSEA was used to calculate the enrichment score of the 18 T-cell related gene sets downloaded from a previous study [53]. The minimal overlap between the gene set and data was fixed at three before the run using the ssGSEA2.0 package.

### Metabolomic analysis using UPLC-MS/MS

Tumor and adjacent non-tumor tissue samples from 17 STX-Hispanic HCC were sent for profiling of global metabolites at the Metabolon Inc. (Durham, NC) facility. Samples were prepared using the automated MicroLab STAR system from the Hamilton Company (Reno, NV). Recovery standards were added before starting the extraction process for quality control purposes. Metabolites were extracted after precipitating proteins with methanol under vigorous agitation for 2 min in Genogrinder 2000 (Glen Mills Inc., Clifton, NJ), followed by centrifugation. The resulting extract was divided into five fractions. Of the four fractions obtained from the extract, two were used for analysis by two separate reverse phase (RP)/UPLC-MS/MS methods with positive ion mode electrospray ionization (ESI), one for analysis by RP/UPLC-MS/MS with negative ion mode ESI, and the last one for analysis by HILIC/UPLC-MS/MS with negative ion mode ESI. All methods utilized an ACQUITY UPLC system (Waters Corp., MA) coupled to a Thermo Scientific Q Exactive high-resolution MS equipped with a heated ESI (HESI-II) source and an Orbitrap mass analyzer. The raw data were extracted, peak-identified and quality control processed using Metabolon’s proprietary hardware and software. Compounds were identified by comparing them with library entries of authenticated standards or recurrent unknown entities. More than 3300 commercially available purified standard compounds have been acquired and registered into LIMS for analysis on all platforms for determination of their analytical characteristics. The area-under-the-curve (AUC) method was applied to quantify peaks. As our present study did not require more than one day of analysis, no normalization was adapted, other than for purposes of data visualization. Detailed methods about UPLC-MS/MS setup and analysis performed at Metabolon Inc. has been described in the article by Ford L. *et al.*[54].

### Pre-processing of metabolomic data

Metabolites with less than 50% missing data from our paired samples were considered for further analyses. The missing data were imputed separately from tumor and adjacent non-tumor samples using Random Forest (RF) algorithm implemented in the R-based missForest (v1.5) package [55] with default parameters. We further removed all drugs and their direct metabolites for the downstream analyses.

### Metabolite set enrichment analysis (MSEA)

We applied the metabolite set enrichment analysis (MSEA) algorithm [56] using the web-based interface of MetaboAnalyst 5.0 (www.metaboanalyst.ca) [57] to identify sub-pathways enriched under the study conditions (tumor and adjacent non-tumor). All metabolites were annotated using the respective Human Metabolome Database (HMDB) IDs. The data were log10 transformed and normalized using the ‘auto-scaling’ option before performing the quantitative enrichment analysis. We used the sub-pathways information provided by Metabolon Inc. as our customized metabolite set library. Analysis was restricted to the metabolite sets containing at least five metabolites.

### Identification of differentially regulated metabolites

Differential regulation of metabolites in tumor vs. paired non-tumor comparison was estimated based on paired sample t-test using log2 transformed data. Metabolites with a minimum 1.5-fold-change and p<0.05 were considered significantly dysregulated in tumors compared to adjacent non-tumor samples.

### GSEA using proteins

We curated a list of gene sets by keywords search for the significantly enriched metabolite sets from GOBP/KEGG/REACTOME entries in the MSigDB (v7.4) collections. A detailed list of 22 curated gene sets has been provided in Table S9. GSEA, using the ranked list of proteins from 15 STX-Hispanic HCC, was conducted with the R package clusterProfiler (v3.18.1) to obtain enrichment scores for the curated gene sets. We performed GSEA using the gene sets with a minimum of 5 and a maximum of 500 genes (proteins), respectively.

### GSEA using metabolites

As the pathway information in our metabolomic data (by Metabolon Inc.) doesn’t include liver function-related hallmark gene sets (MSigDB), we curated a knowledge-based collection of metabolite sets (Table S10). We ranked ∼ 700 metabolites for (a) overall STX-Hispanic HCC using average log2 (fold-change) and (b) each patient using log2 (fold-change) metric from tumor vs. paired non-tumor comparison. Enrichment of curated metabolite sets from ranked metabolites was obtained using GSEA and ssGSEA with clusterProfiler and ssGSEA2.0, respectively.

### Serum lipidomic analysis using multi-dimensional mass spectrometry-based shotgun lipidomics

Lipid species were analyzed using multidimensional mass spectrometry-based shotgun lipidomic analysis [58]. In brief, each serum sample (100 µl) was transferred to a disposable glass culture test tube. A premixture of lipid internal standards (IS) was added prior to conducting lipid extraction for quantification of the targeted lipid species. Lipid extraction was performed using a modified Bligh and Dyer procedure [59], and each lipid extract was reconstituted in chloroform:methanol (1:1, v:v) at a volume of 2 µl/µl serum.

For shotgun lipidomics, lipid extract was further diluted to a final concentration of ∼500 fmol total lipids per microliter. Mass spectrometric analysis was performed on a triple quadrupole mass spectrometer (TSQ Altis, Thermo Fisher Scientific, San Jose, CA) and a Q Exactive mass spectrometer (Thermo Scientific, San Jose, CA), both of which were equipped with an automated nanospray device (TriVersa NanoMate, Advion Bioscience Ltd., Ithaca, NY) as described [60]. Identification and quantification of lipid species were performed using an automated software program [61]. Data processing (e.g., ion peak selection, baseline correction, data transfer, peak intensity comparison and quantitation) was performed as described [61]. The result was normalized to the serum volume (nmol lipids/mL serum).

### Statistical analyses for serum lipidomic data

Differences in total lipid concentrations of each lipid category between HCC patients and non-HCC individuals were assessed using the Wilcoxon test in R package (v4.0.4). Differential overall lipid profiles between HCC and non-HCC, and between STX−Hispanic and STX−non-Hispanic were identified by Wilcoxon test using log2 transformed average lipid concentrations.

## Results

### Patient samples and ancestry profiles

Clinical information and tissue samples (tumor and adjacent non-tumor) were collected from 31 Hispanic HCC patients in South Texas (STX-Hispanic) (Fig. 1A and Table S1). We only included patients without evidence of any regional or distant metastases during the sample collections. Among the 31 patients, the majority were male (64.5%), HBV/HCV positive (54.8%), diabetic (54.8%), and obese (BMI ≥ 30)/overweight (25.0 ≤ BMI < 30) (64.5%). Besides, ∼52% of the tumors were high-grade (grades 3 and 4), and ∼29% were high-stage (pathological stages T3 and T4). Whole exome sequencing (WES) was performed on 27 (discovery cohort) of the 31 paired tumor-nontumor samples to identify somatic mutations. We collected another 38 tumor-only samples (validation cohort) from STX-Hispanic HCC to validate the mutation frequencies observed in our discovery cohort patients. RNA sequencing was performed on 13 paired tumor-nontumor samples, which met quality control requirement for the transcriptomic analysis. Mass spectrometry (MS)-based proteomic and metabolomic data were generated for 15 and 17 paired tumor-nontumor samples, respectively (Fig. 1A).

**Figure 1:**
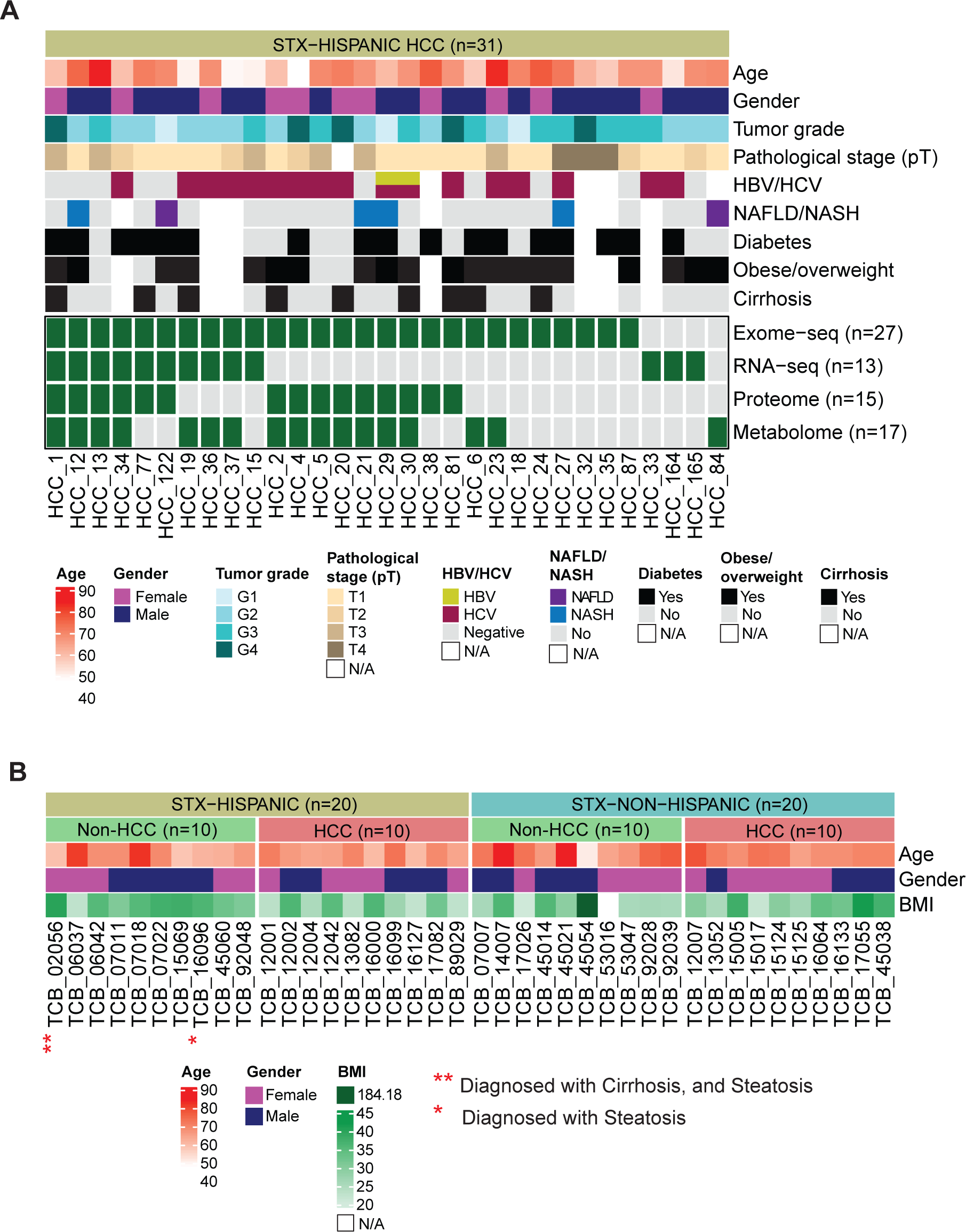
Overview of demographic and clinical information of HCC patients and sequencing data. (A) Multi-omics data modalities for paired tumor and non-tumor tissues from STX-Hispanic HCC are shown in different rows. Each column indicates one patient. (B) The clinical characteristics of 40 samples screened for serum lipid profiles.

Based on a panel of in-house curated ancestry informative markers (AIMs) from WES data on the non-tumor samples of the 27 HCC (discovery cohort) patients [21], we confirmed genetic admixture in our cohort of STX-Hispanic patients. They all showed a genetic admixture between European (EUR) and East Asian (EAS) and, to a lesser extent, African (AFR) ancestral traits (Fig. S1).

We also performed mass spectrometry-based shotgun lipidomics analysis of serum samples collected from 20 STX-HCC and 20 STX-non-HCC individuals with equal distribution of Hispanics and non-Hispanics (Fig. 1B). We included individuals with no evidence of (a) HBV or HCV infections; (b) associated NAFLD/NASH; and (c) heavy alcohol intake, to avoid potential non-HCC-specific alterations in the serum lipidomic data. Their liver had no cirrhosis except in one STX-Hispanic non-HCC patient (Fig. 1B and Table S2). A written informed consent was provided by all study participants.

### Patient demographic and clinical parameters do not confound omics data in STX-Hispanic HCC

To determine whether the observed tumor-associated alterations of the omics data might be confounded by patient demographic and/or clinical features, we performed multivariate regression analysis and found that the frequently mutated genes reported in STX-Hispanic HCC (Fig. 2A) are not significantly (adjusted p-value=1) associated with any of the patient demographic and clinical features including age, sex, viral status, NAFLD/NASH, cirrhosis, obesity, and diabetes. We also investigated the effect of age, sex, viral status, obesity, and diabetes status on mRNA/protein/metabolite levels in our STX-Hispanic HCC datasets, using multivariate regression models with mRNAs/proteins/metabolites-based log2 (fold-change) in tumor vs. adjacent non-tumor comparison as the outcome. Only *c19orf79* (also known as *PET100*) had a significant negative association with age (FDR=0.016) and a positive association with diabetes status (FDR=0.028) in the mRNA data. The gene was not significantly differentially expressed in tumors. The log2 (fold-change) of proteins and metabolites showed no significant association with those demographic and clinical variables. Thus, the log2 (fold-change) between tumor and adjacent non-tumor tissue is a feasible measurement for investigating HCC-specific alterations. The lack of association of the common patient demographic and clinical parameters with genetic and epigenetic alterations led us include HCC tumors with different patient demographics and etiology for multi-omic studies.

**Figure 2:**
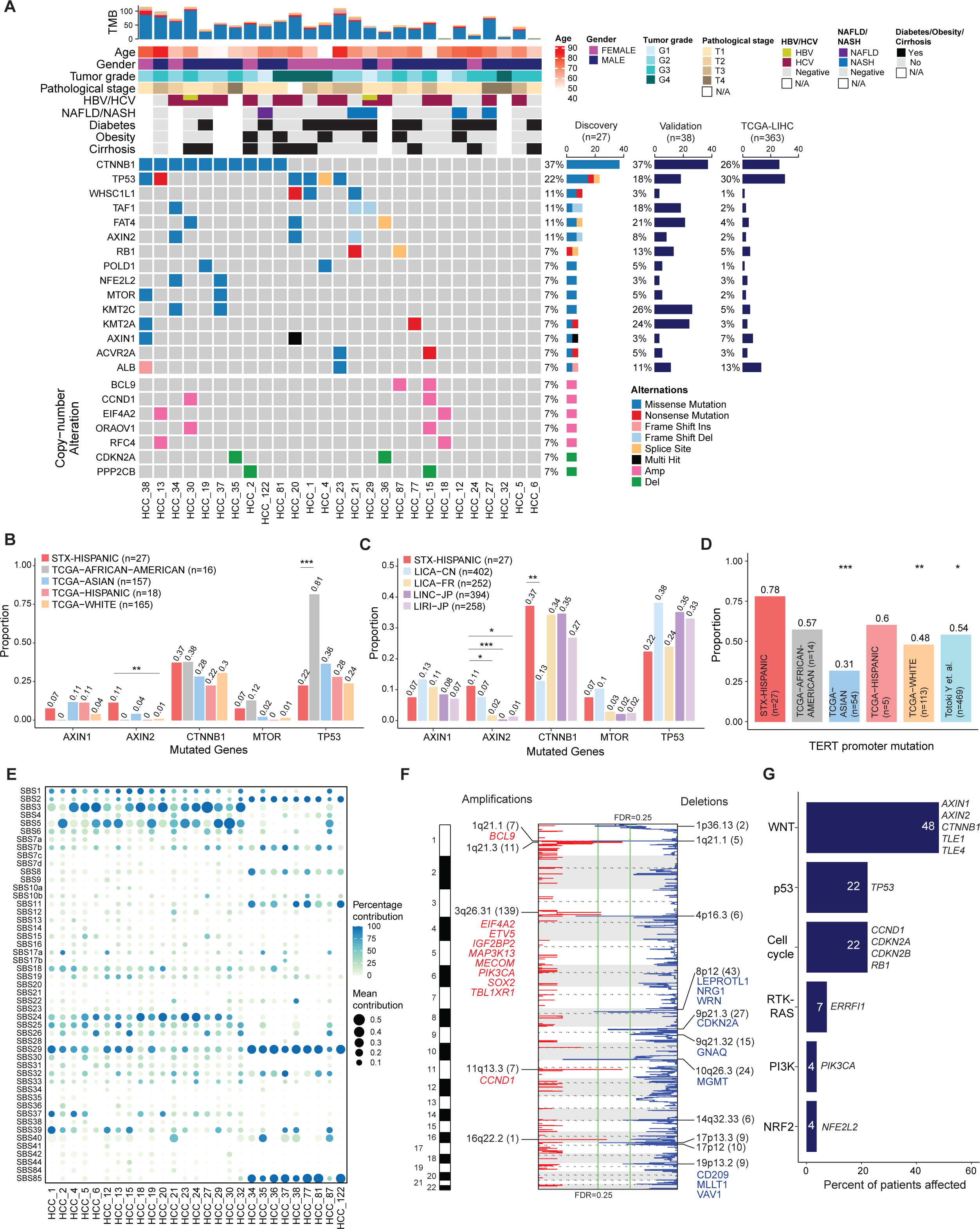
The mutational landscape of HCC in STX-Hispanic patients. (A) Genetic profile and associated clinicopathological characteristics of 27 STX-Hispanic HCC. Bar plots on the right indicate mutation frequencies. Left: 27 discovery samples; middle: 38 validation samples; right: 363 TCGA-LIHC samples. (B-C) Comparison of mutation frequencies of five recurrently mutated genes between STX-Hispanic HCC and TCGA-LIHC or ICGC HCC cohorts of different ethnicities. (D) Comparison of *TERT* promoter mutation frequencies between HCC from different ethnicities. For B-D, p-values are calculated using Fisher’s exact test. Significance level: *, p<0.05; **, p<0.001; ***, p<0.001). (E) Mutational signatures in 27 HCC tumors. The size of each dot indicates the average contribution of that signature (in the iterations where contribution was higher than 0). Color of dots are representative of percentage of iterations in which the signature is found (contribution > 0). (F) Recurrent copy number amplifications and deletions across 27 STX-Hispanic HCC by GISTIC 2.0. Annotated peaks have an FDR<0.25. The numbers in parentheses indicate the number of genes encompassed in the peak. Cancer genes from CGC (COSMIC) are annotated for each peak. Red, amplification; Blue, deletion. (G) Alteration frequency of oncogenic signaling pathways in STX-Hispanic HCC.

### Genetic alterations and characteristics of STX-Hispanic HCC

We identified 1,528 somatic variants in the 27 HCCs from the discovery cohort using VarScan2 and MuTect2. The median non-silent mutation burden was 1.06 mutations/Mb (range: 0.04-2.28). Ninety-five genes were mutated in at least two (∼7%) patients. Eighty-four of these 95 genes were also found mutated in one or more patients in the validation cohort. Fig. 2A shows genes with somatic mutation frequency ≥ 7% that are either significantly mutated (MutSigCV, p<0.05) (n=9) or annotated in the cancer gene census (CGC, COSMIC) (n=5), or among the significantly mutated genes reported by the TCGA-LIHC study (n=1). All these genes were expressed (average FPKM > 0.5). The somatic mutation frequencies of these genes were overall consistent between our cohort and the TCGA-LIHC cohort (Fig. 2A, Fig. S2a). However, higher mutation frequencies were observed for *WHSC1L1* (11.1% vs. 0.6%, p=0.00271), *TAF1* (11.1% vs. 1.9%, p=0.0257), and *AXIN2* (11.1% vs. 1.9%, p=0.0257) (Fig. S2a). Considering ethnic groups, *AXIN2* mutation frequency was significantly higher in STX-Hispanic HCCs than Whites (11.1% vs. 0.6%, p=0.00912) in the TCGA-LIHC cohort [12] and HCC cohorts in Japan (11.1% vs. 0.3%, p=0.00091 for LINC-JP; 11.1% vs. 1.2%, p=0.0126 for LIRI-JP) and France (11.1% vs. 1.6%, p=0.0218) [30].

To confirm all *AXIN2* mutations observed in our patient cohort, we performed DuplexSeq-based targeted DNA sequencing. We validated all three mutations detected in *AXIN2* (Fig. S2b, Table S3). We also validated mutations in *MTOR* (Fig. S2b, Table S3), a well-known proto-oncogene, which was relatively more frequent in STX-Hispanic HCCs than in other ethnic groups (7.4% vs. 1.9% in TCGA-Asian or 1.2% in TCGA-White), even though not statistically significant (Fig. 2B, 2C). Other notable observations were a lower *TP53* mutation frequency and a higher *CTNNB1* mutation frequency in the STX-Hispanic cohort than the African−Americans from the TCGA (p=0.00032) (Fig. 2B) and HCC cohort in China (p=0.00195) (Fig. 2C), respectively.

We sequenced two independent activating mutations located at −124 (C228T) and −146 (C250T) base-pairs upstream of the transcription start site frequently seen in the TERT promoter in many HCC cohorts including the TCGA-LIHC [12]. We identified a significantly higher rate of TERT promoter mutation (primarily C228T) in the STX-Hispanic HCC cohort than that in the TCGA-LIHC White (77.8% vs. 47.8%, p=0.00535) and Asian patients (77.8% vs. 31.5%, p=0.00012), and another study population from Totoki et al. (77.8% vs. 53.9%, p=0.0167) [62] (Fig. 2D). Thus, mutations of the Wnt pathway genes and TERT promoter appear to play a more dominant role in hepatic carcinogenesis in the STX-Hispanic HCC cohort than in other ethnic cohorts.

The mutational signature using the 96 different contexts of single base substitution (SBS) revealed similarities and differences among STX-Hispanic HCCs (Fig. S2c). Considering SBS signatures found in > 50% bootstrap iterations, eight were frequently identified, at least in one-third of our patient cohort. The eight SBS signatures and their frequencies in our HCC cohort were SBS1 (5-methylcytosine; n=19), SBS29 (tobacco chewing; n=19), SBS2 (APOBEC; n=14), SBS24 (aflatoxin exposure; n=14), SBS3 (homologous recombination deficiency; n=13), SBS5 (clock-like; n=12), SBS7b (UV exposure; n=11), and SBS39 (n=9) (Fig. 2E). Of these signatures, SBS1, SBS24, SBS29, and SBS5 were previously found in HCC by ICGC pan-cancer consortium (PCAWG) [33]. However, SBS2, SBS3, and SBS7b were not frequently detected in HCC by PCAWG [33].

Somatic copy number alterations (SCNAs) were identified using VarScan2 and GISTIC2.0 algorithm from WES. We identified 5 focal amplification peaks and 11 deletion peaks (q-value<0.25). Nineteen genes from these regions were cataloged in the CGC (COSMIC) (Fig. 2F), including oncogenes *BCL9* (1q21.1), *PIK3CA*, *MAP3K13*, *TBL1XR1* (all from 3q26.31), and *CCND1* (11q13.3) within the amplification peaks and tumor suppressor genes *CDKN2A* (9p21.3) and *MGMT* (10q26.3) within the deletion peaks (Fig. 2F). The recurrent gain of *BCL9*, *CCND1*, *ORAOV1*, and loss of *CDKN2A* shown in Fig. 2A have been reported in other HCC cohorts [12,63,64]. We also observed recurrent gain of *EIF4A2* and *RFC4*, and loss of *PPP2CB* in our STX-Hispanic cohort (Fig. 2A). SCNAs of those genes were also significantly positively correlated (Pearson’s correlation; r>0, FDR<0.01) with their mRNA expression (FPKM) levels. We observed no significant differences in the frequency of GISTIC2.0 high-level copy number amplifications/deletions between STX-Hispanic HCC tumors and other cohorts (Fig. S2d) in cBioPortal [65].

Inspecting the set of ten well-recognized oncogenic signaling pathways [35] using potentially oncogenic (OncoKB curated) somatic non-silent mutations and SCNAs, our STX-Hispanic HCC cohort showed recurrent alterations in WNT, p53, Cell cycle, RTK/RAS pathways. The first three pathways were altered in more than 22 percent of patients (Fig. 2G).

### Integrative analysis of transcriptomic, proteomic, and genomic profiles of STX-Hispanic HCC

We identified 1,188 differentially expressed protein-coding genes (DEGs) between 13 pairs of tumor and non-tumor samples (Fig. 3A and Table S4). We also identified nine fusion events (Table S5). Among the fusion genes, *HSD17B2* was also significantly downregulated in the HCC tumors (Table S4). Ninety-two genes were found to undergo one or more significant alternative splicing (AS) in the tumors (Fig. S3a and Table S6). Of the 92 genes, nine (*C10orf116*, *CYP4A11*, *ECHDC2*, *FAHD2A*, *NDRG2*, *RGN*, *SIGIRR*, *GCH1,* and *ZGPAT*) were significantly downregulated in the HCC tumors. Skipped exon (SE) event was seen in the first seven genes and alternative 3′ splice site (A3SS) event in *ZGPAT*. Co-occurrence of SE and A3SS events were seen in *GCH1*. The significance of these fusion and AS events in the tumors remains to be investigated.

**Figure 3:**
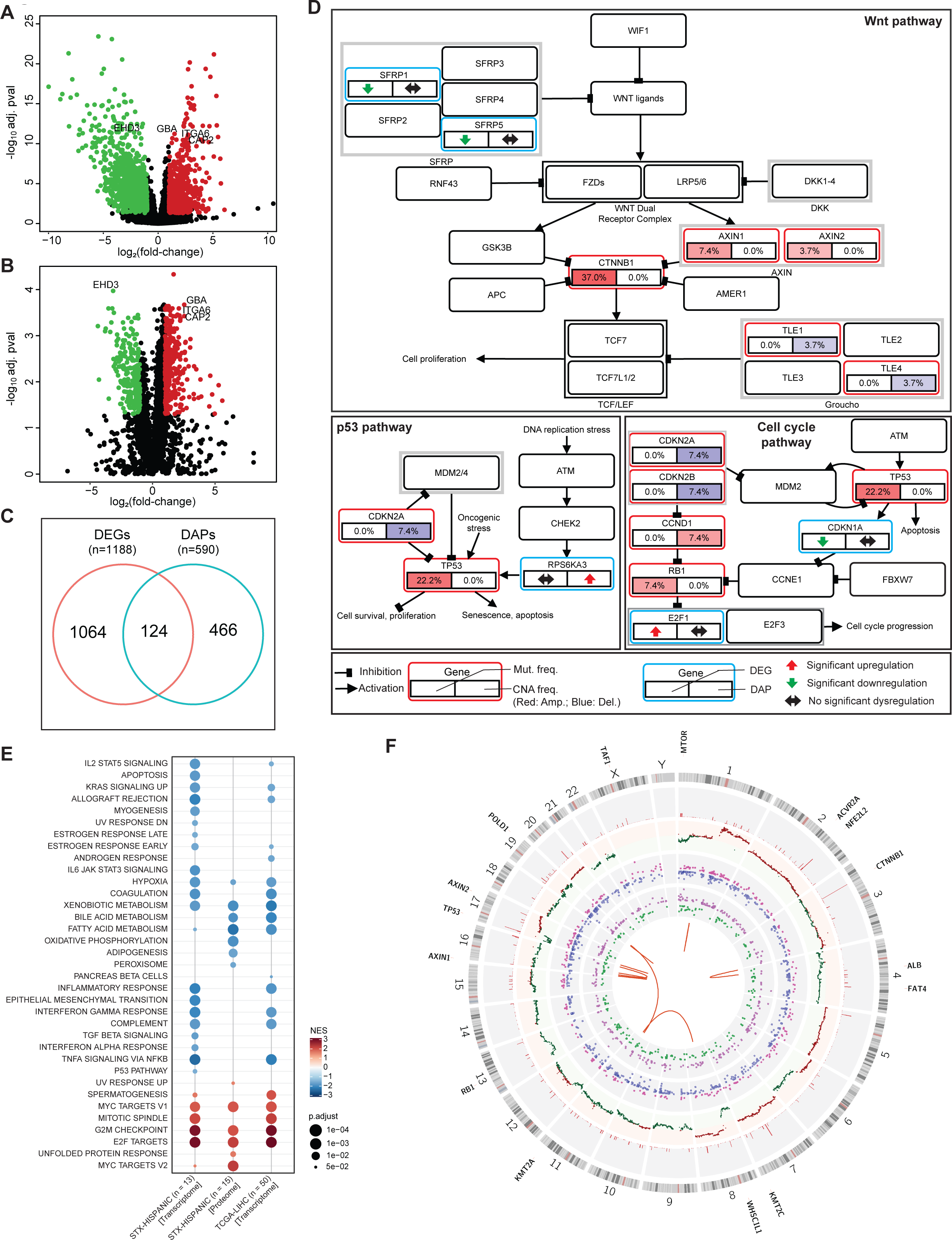
The integrated multi-omics profiling of STX-Hispanic HCC. Volcano plots illustrate (A) DEGs and (B) DAPs between tumor and adjacent non-tumor samples. Symbols indicates common genes and proteins among 10% most significant DEGs and DAPs, respectively. Red, up regulated; Green, down regulated. (C) The intersection of DEGs and DAPs. (D) Schematic of top three recurrently altered oncogenic signaling pathways in STX-Hispanic HCC. Genes with somatic mutations and copy number alterations are indicated in red boxes. Color intensity indicates the frequency of activating alterations (red) and inactivating alterations (blue). The frequencies were calculated based on the discovery cohort (n=27). The cyan boxes are indicative of DEGs and DAPs. Up- and down-regulated DEGs (left) / DAPs (right) are shown in red and green arrows, respectively. (E) Hallmark gene sets significantly enriched in STX-Hispanic HCC and TCGA-LIHC. Red dots, positive enrichments. Blue dots, negative enrichments. (F) Circos plot shows genomic, transcriptomic, and proteomic alterations in STX-Hispanic HCC. From outside to inside, the tracks depict the human genome (hg19) ideogram, frequency (number) of genetic alteration, GISTIC 2.0 copy-number amplification (red) and deletion (green), log2 (fold-change) of upregulated (pink) and downregulated (blue) DEGs, log2 (fold-change) of increased (purple) and decreased (green) DAPs, fusion genes (orange ribbons). Recurrently mutated genes are noted outside the ideogram.

Using paired sample t-test, we identified 590 differentially abundant proteins (DAPs) between tumors and non-tumor controls (Fold-change ≥ 2, FDR<0.05) (Fig. 3B and Table S7). Overall, 124 genes were differentially expressed (DEGs) in parallel with their corresponding proteins (DAPs) (Fig. 3C), which is 33.3% (i.e. 124 of 372 common proteins) or 21.9% (i.e. 124 of 566 common genes) of total number of DEGs or DAPs, respectively. Many of the DEGs and DAPs were constituents of various oncogenic pathways including the top ranked Wnt, p53, and cell cycle pathways that were enriched with somatic alterations (Fig. 3D).

To further examine the correlation between mRNA expression and protein abundance, we performed Pearson’s correlation analysis for each of 5,460 gene-protein pairs detected from both transcriptomic and proteomic analyses in the tumor and adjacent non-tumor tissues of the six cases. The majority (∼76.4%) of the gene-protein pairs showed positive correlation (Fig. S3b, left panel) indicating a general concordance between mRNA and protein abundances. Consistently, gene set enrichment analysis (GSEA) with the 50 hallmark gene sets showed that seven of the 13 (54%) significantly enriched hallmark gene sets using the proteome data were also significantly enriched using the transcriptome data of the STX-Hispanic cohort (Fig. 3E). Mean of the pairwise correlation coefficients between mRNAs and proteins originated from the 2,123 genes of the 50 hallmark gene sets was higher (0.37 vs. 0.25) than that from the 3,337 non-hallmark genes (Fig. S3b, right panel), suggesting a better concordance between mRNA and protein abundances of the hallmark genes. While the overall enrichment pattern is similar between STX-Hispanic and TCGA-LIHC transcriptome data (Fig. S3c), STX-Hispanic cohort showed more negatively enriched gene sets than the TCGA-LIHC cohort, including apoptosis, IL6 Jak Stat3 signaling, and epithelial mesenchymal transition (Fig. 3E).

Chromosomal distributions of all genomic, transcriptomic, and proteomic alterations from our Hispanic HCC cohort are shown in the Fig. 3F. Integrated genomic and proteomic data revealed overall higher (but not significant) upregulation of mutated CTNNB1 protein (n=7) compared to wild-type protein (n=8) in tumors of STX-Hispanic HCC (Fig. S3d). Abundance of other five most recurrently mutated genes (with mutation frequency > 10%) could not be identified from STX-Hispanic HCC proteomic data.

### Proteo-transcriptomic enrichment analyses of HCC identify two clusters with different enrichments of hallmark gene signatures

Unsupervised clustering of STX-Hispanic HCC (transcriptome- and proteome-based) and TCGA-LIHC (transcriptome-based) cohorts independently separated them to two clusters (labelled as HM-1 and HM-2). Based on function, we categorized the hallmark gene sets to four groups as immune related, cell-cycle related, liver function (fn)-related, and others (Fig. 4A). We further confirmed our two clusters using principal component analysis (PCA) of transcriptome-based NES from STX-Hispanic and TCGA-LIHC combined dataset (Fig. S4a). As anticipated, the six STX-Hispanic cases with both transcriptomic and proteomic data were segregated into the same cluster with either the transcriptomic or proteomic data (Fig. 4A), suggesting again the overall concordance between protein abundance and mRNA expression.

**Figure 4:**
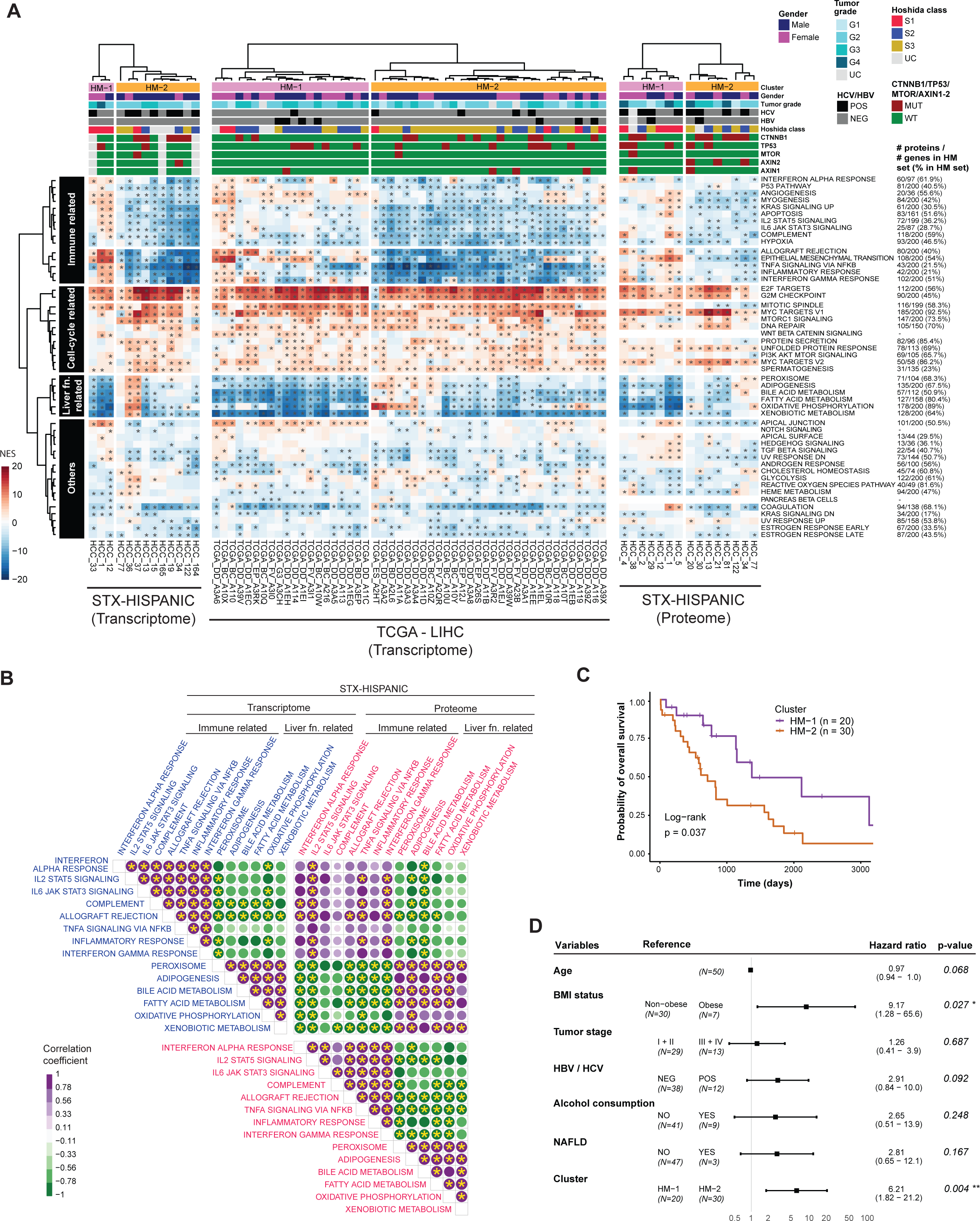
Proteomics and transcriptomics-based classification of HCC. (A) Left panel: HM-1 and HM-2 subtypes from unsupervised hierarchical clustering of pathway activities in STX-Hispanic HCCs. Middle: TCGA-LIHC. Right: STX-Hispanic proteomics cohort. Pathway activity is based on NES from ssGSEA analysis. Asterisks indicate significant (FDR<0.05) enrichments. (B) Comparison between enrichment correlations of selected immune and liver-function-related hallmark gene sets using NES from transcriptome and proteome data of STX-Hispanic HCC (based on 6 HCC with both datasets). An asterisk indicates significant (Pearson’s correlation; p<0.05) association between a pair of gene sets. (C) Kaplan-Meier curve for overall survival in 50 TCGA-LIHC stratified by two clusters – HM-1 and HM-2. (D) Multivariable Cox regression analysis of overall survival in the TCGA cohort (n=50) controlling for other clinicopathological factors.

Cell-cycle-related hallmark gene sets were almost uniformly positively enriched in each patient. HM-1 cases showed general positive NESs of the Immune related hallmark gene sets, whereas HM-2 had negative NESs. The overall negative enrichment of the immune-related gene sets (Fig. 3E) was apparently due to a larger number of HM-2 cases with a negative NES (Fig. 4A). Compared with HM-2 cases, HM-1 cases showed more negative NESs of the liver fn-related hallmark gene sets (Fig. 4A), suggesting a negative correlation between immune function and liver function in HCC tumors. We further confirmed this negative correlation in both transcriptome and proteome data (Fig. 4B).

Because transcriptome- and proteome-based clustering produced consistent results in our cohort, we were able to divide 22 patients into HM-1 (n = 8) or HM-2 (n = 14) cluster based on either transcriptome or proteome data. We found HM-2 cluster had a greater percentage (50% vs. 14%) of Hoshida Class S3 patients [50] and more patients (83% vs. 29%) with genomic alterations in Wnt pathway than the HM-1 cluster (Fig. S4b). Kaplan-Meier analysis of the 50 TCGA-LIHC cohort revealed a significantly worse (log-rank test, p=0.037) overall survival (OS) of HM-2 patients than HM-1 patients (Fig. 4C). Multivariate Cox regression analysis controlling for BMI, age, tumor stage, viral (HBV / HCV) infection, alcohol consumption, and NAFLD (nonalcoholic fatty liver disease) confirmed the independent prognostic significance of the cluster (p=0.004; Fig. 4D).

### Differences between HM-1 and HM-2 HCC in tumor immune microenvironment

The substantial differences in the enrichments of immune-related hallmark gene sets between HM-1 and HM-2 tumors led us to investigate the tumor immune microenvironment of these two HCC subtypes. Regardless of ethnicity, we found significantly (Wilcoxon test, p<0.05) higher immune and stromal scores, and significantly lower tumor purity in HM-1 tumors than HM-2 tumors (Fig. 5A). To further characterize the tumor immune microenvironment, we used the CIBERSORT absolute method to estimate the abundance of 22 immune cell types in the tumor and adjacent non-tumor tissues. In the TCGA-LIHC cohort, the abundance of all macrophages (i.e., M0, M1, and M2), activated NK cells, and resting mast cells were significantly (Welch’s t-test, p<0.05) higher in HM-1 tumors than HM-2 tumors (Fig. S5a). Similarly, various types of T cells including CD8, CD4 memory resting, follicular helper, and regulatory (Tregs) cells were also significantly more abundant in HM-1 tumors. In the STX-Hispanic cohort, we also observed a similar increase of these immune cells in HM-1 tumors (Fig. S5a). We observed an opposite trend of infiltrated CD8 T-cell abundance in HM-1 and HM-2 tumors when compared with paired adjacent non-tumor samples of both STX-Hispanic and TCGA-LIHC cohorts (Fig. S5b).

**Figure 5:**
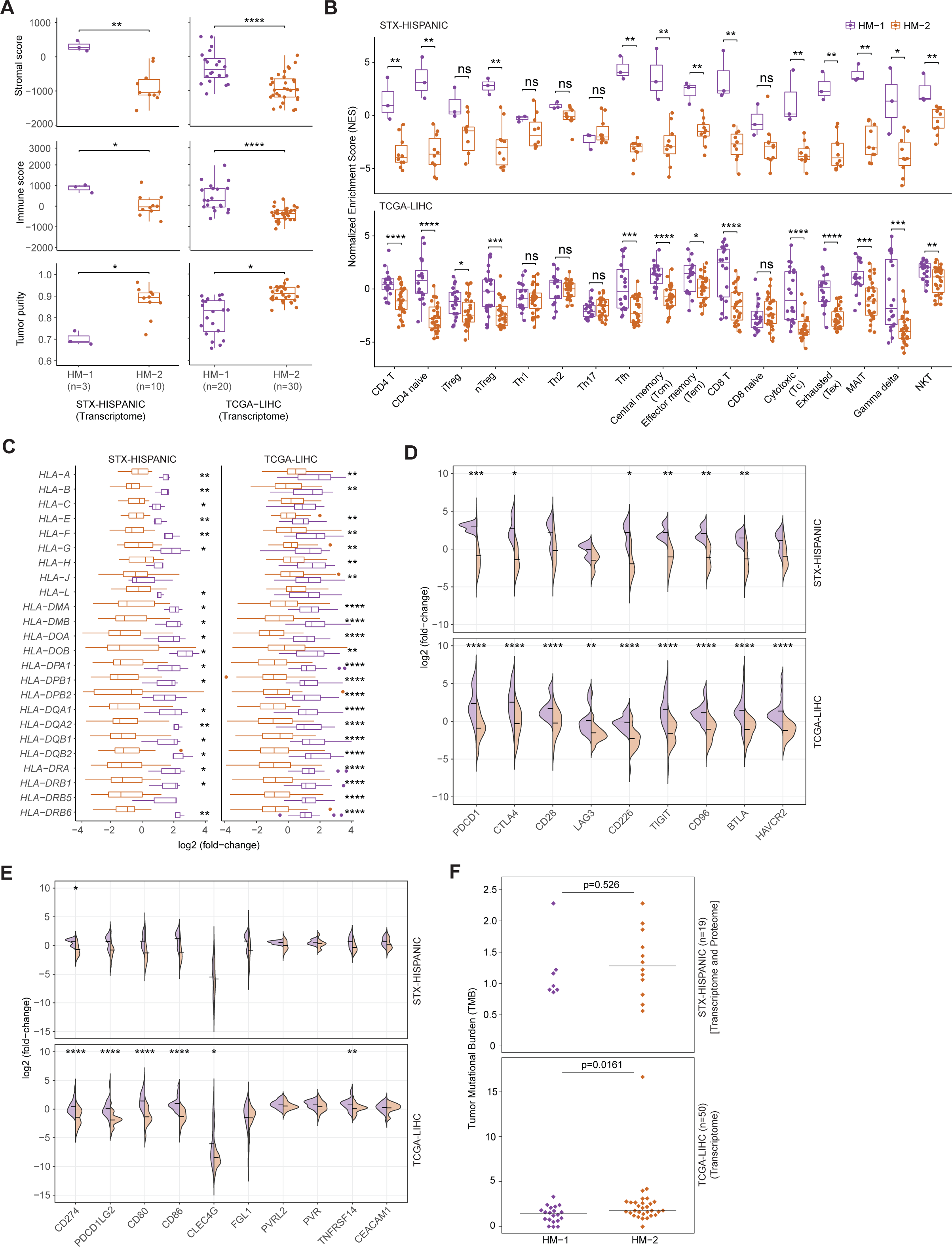
Immune-cell profiling of HCC identifies significant differences between the two patient clusters. (A) Comparison of the stromal score, immune score (immune cell infiltration levels), and tumor purity between HCC clusters in STX-Hispanic and TCGA cohorts. (B) HCC clusters-wise differences in the enrichment of T-cell-related gene sets (n=17). (C) Comparison of log2 (fold-change) of HLA gene expression (tumor vs. paired non-tumor) between the two HCC clusters. (D) Differential expression of immune checkpoint receptors and between the clusters. (E) Differential expression of immune checkpoint ligands between the clusters. (F) Difference in Tumor Mutational Burden between HM-1 and HM-2 clusters. Upper, STX-Hispanics; Lower, TCGA-LIHC. Statistical significance for all comparisons in A-C and F was determined using two-sided Wilcoxon rank-sum tests. The p-values for D and E were calculated using unpaired two-tailed t-test.

We applied the ssGSEA algorithm to investigate the expression activities of T-cells [53] in the STX-Hispanic and TCGA HCC cohorts. The NESs for the majority T cells including CD4, CD8, cytotoxic, exhausted, and natural killer T cells were significantly higher in HM-1 tumors than HM-2 tumors of both study cohorts (Fig. 5B). Considering relative expression of human leukocyte antigen (HLA) [also known as major histocompatibility complex (MHC)] genes in tumor vs. non-tumor tissues, the majority were upregulated (median log2 fold-change > 0) in HM-1 tumors and downregulated (median log2 fold-change < 0) in HM-2 tumors. Moreover, a significant difference in fold-changes of the majority of MHC genes was seen between HM-1 and HM-2 of both HCC cohorts (Fig. 5C). The significant enrichment of CD4+, CD8+, cytotoxic T-cells, and higher MHC expression in the HM-1 tumors suggest enhanced host immune defense against these tumors. In contrast, HM-1 tumors showed increased enrichment of exhausted T cells, suggesting they may be more responsive to immune checkpoint blockade. Fig. 5D shows the expression of immune checkpoint (IC) receptors mainly upregulated in the tumors over paired non-tumor tissues and the mean log2 fold-changes were significantly higher in the HM-1 subtype than those in the HM-2 subtype of both STX-Hispanic and TCGA-LIHC cohorts. Similarly, the mean fold-changes of several IC ligands including *CD274* (*PD-L1*), *PDCD1LG2* (*PD-L2*), *CD80*, *CD86*, and *TNFRSF14* (*HVEM*) were significantly higher in the HM-1 subtype than the HM-2 subtype of TCGA-LIHC cohort. A similar pattern was also observed in STX-Hispanic cohort (Fig. 5E).

Pearson correlation analysis revealed a significant positive association between CD8+ T-cell abundances (CIBERSORT) and the expression (log2 FPKM) of IC receptors in both STX-Hispanic (Fig. S5c) and TCGA-LIHC (Fig. S5d) tumors, suggesting that the receptors were likely expressed by CD8 T cells. The significant positive correlation of the expression of some IC receptors with the expression of their ligands, including PD-1/PD-L1, PD-1/PD-L2, CTLA4/CD80, CTLA4/CD86, and CD28/CD86, in both STX-Hispanic (Fig. S5e) and TCGA-LIHC (Fig. S5f) tumors suggests that these IC ligands were mainly expressed by CD8 T cells. Other IC ligands, mostly showing a non-significant correlation with their receptors, are probably expressed by different cell types. Besides, positive correlation between BTLA (receptor) and HVEM (ligand) were nearly significant in both STX-Hispanic HCC (p=0.081, Fig. S5e) and TCGA-LIHC (p=0.059, Fig. S5f).

Taken together, these data suggest that while HM-1 tumors appear “immune hot” with increased T cells, some of which are apparently in an exhaustion state with activated IC signaling. As such, HM-1 tumors should be more responsive to IC inhibition (ICI) therapy. On the other hand, we found that tumor mutational burden (TMB), an indicator of response to ICI therapy, was slightly lower in HM-1 tumors than in HM-2 tumors (Fig. 5F). Thus, while the IC inhibitor atezolizumab plus the angiogenesis inhibitor bevacizumab is currently the frontline therapy for advanced HCC, whether HM-1 tumors are more responsive to the therapy than HM-2 tumors remains to be investigated.

### Significant alterations of metabolic pathways in STX-Hispanic HCC

Using quantified metabolites (n=704), partial least squares-discriminant analysis (PLS-DA) was performed on the 17 paired tumor-nontumor samples (Fig. 6A). The PLS-DA score plot result shows a clear separation between the metabolite levels in the tumor and adjacent non-tumor tissues accounting for 18.5% of the total variability observed in the data set. Eleven metabolic pathways were significantly (p<0.05) enriched in the tumors of HCC cohort on quantitative metabolite set enrichment analysis (MSEA) [56] (Fig. 6B). Differential regulation analysis using paired sample t-test revealed 131 metabolites with significant (p<0.05) dysregulation by ≥ 1.5-fold in the tumors compared to paired non-tumor tissues (Table S8). Significantly dysregulated metabolites associated with the 11 functionally enriched pathways are shown in Fig. 6B.

**Figure 6:**
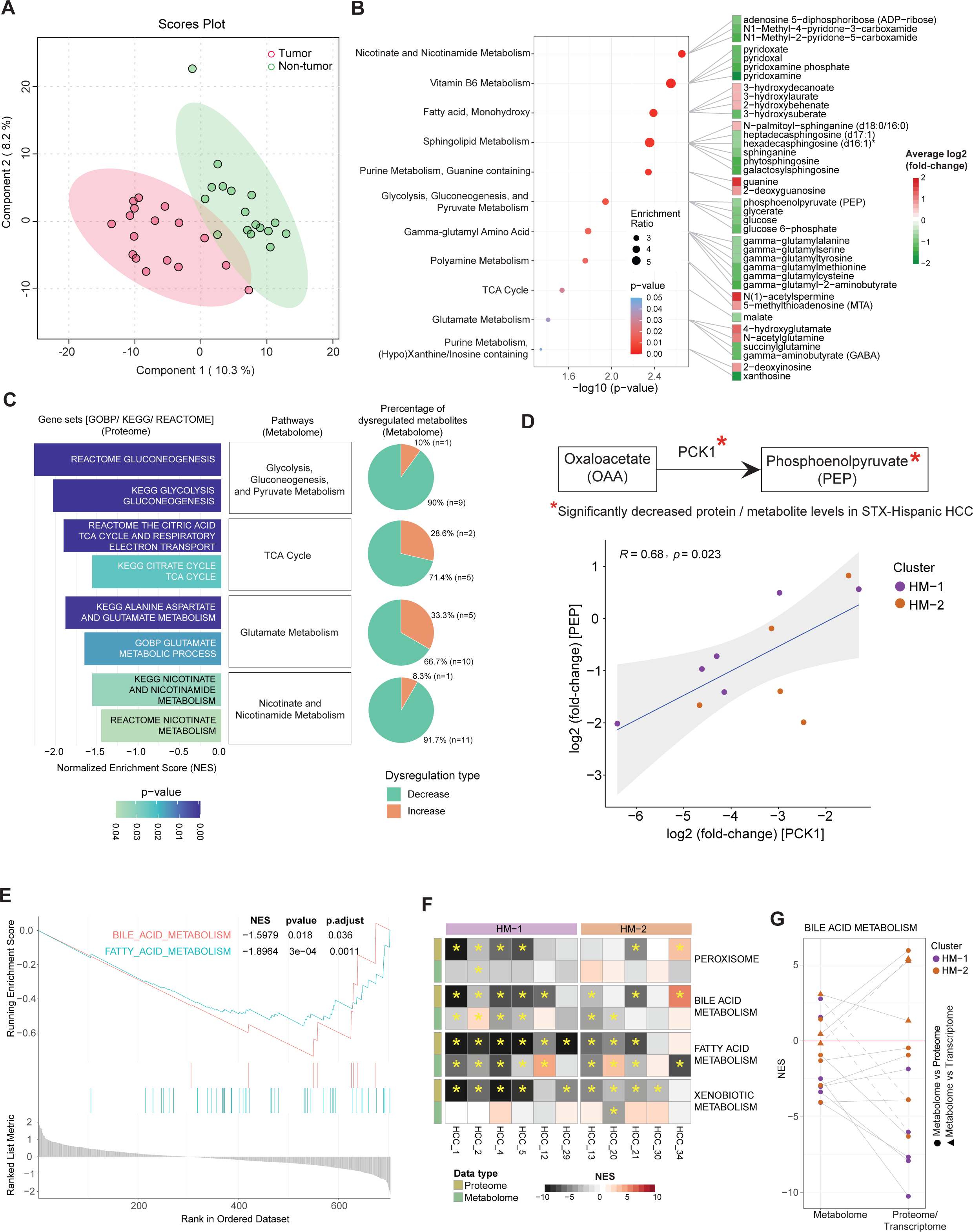
Tissue metabolomic profiles of HCC in STX-Hispanics. (A) Score plot for PLS-DA analysis of tumor (red) and adjacent non-tumor (green) tissue samples derived from STX-Hispanic HCC. (B) Eleven significantly enriched pathways in HCC tumors (p<0.05). The right panel indicates log2 (fold-change) of significantly enriched (paired sample t-test, p<0.05; and ≥ 1.5 fold-change) metabolites against each pathway. (C) Eight significantly negatively enriched GOBP/KEGG/REACTOME pathways (p<0.05) were obtained from GSEA analysis using protein abundance in STX-Hispanic HCC (left). Pie-chart representation of the directional dysregulation of metabolites (tumor vs. paired non-tumor comparison) from each metabolomic pathway is shown on the right. (D) A scatterplot of log2 (fold-change) between PCK1 (protein) and PEP (metabolite) from STX-Hispanic HCC (n=11) shows a significant correlation. The interaction between the two is shown above. (E) Enrichment plots of two significantly negatively enriched pathways curated using selected metabolites. (F) Comparison of NES between hallmark gene sets and curated metabolite sets from ssGSEA analyses using proteome and metabolome data, respectively. The asterisk indicates significant enrichments (FDR<0.05). (G) Comparison of enrichment (NES) trends for bile acid metabolism between proteomic/transcriptomic and metabolomic data sets in STX-Hispanic HCC (n=14). Solid lines indicate concordance and dashed lines indicate discordance in the directionality of enrichments.

Since altered metabolites and metabolic pathways are mainly caused by altered protein/enzyme abundances, we used proteome-based GSEA to find enrichment of gene sets (Table S9) that were functionally similar to the 11 enriched metabolic pathways. Eight GOBP/KEGG/REACTOME-derived gene sets corresponding to four of the 11 metabolic pathways were significantly negatively enriched in STX-Hispanic HCC (Fig. 6C, left). Metabolites associated with the four negatively enriched metabolic pathways were found predominantly decreased in HCC tumors (Fig. 6C, right). Therefore, integrated analysis of our proteome and metabolome data of STX-Hispanic HCC revealed potential negative enrichments of (a) glycolysis and gluconeogenesis, (b) TCA cycle, (c) glutamate metabolism, and (d) nicotinate and nicotinamide metabolism processes. Furthermore, we calculated the association between tumor vs. non-tumor log2 (fold-change) of substrates or products (considering only significantly differentially dysregulated metabolites from these four pathways) and corresponding enzymes (considering only DEPs) for 11 STX-Hispanic HCC with both proteome and metabolome data. We observed significant association (Pearson’s correlation, r = 0.68, p=0.023) of log2 (fold-change) between phosphoenolpyruvate (PEP) and phosphoenolpyruvate carboxykinase 1 (PCK1) (Fig. 6D). Thus, PCK1—a rate-limiting enzyme in the gluconeogenic pathway [66]— and PEP could serve as putative biomarkers for the identification and treatment of HCC.

We next investigated whether the significant negative enrichments of the four liver function-related hallmark gene sets (bile acid metabolism, fatty acid metabolism, peroxisome, and xenobiotic metabolism) from GSEA of our proteome data (Fig. 3E) are associated with differences in relative abundance of corresponding metabolites between tumor and non-tumor tissues. We performed enrichment analyses of a knowledge-based collection of curated metabolite sets (Table S10) for overall GSEA or ssGSEA. Bile acids and fatty acids were significantly (corrected p-value=0.03 and 0.0004) negatively enriched (NES = −1.595 and −1.872) in STX-Hispanic HCC tumors (Fig. 6E). Comparison of the NES between proteome data and metabolome data from ssGSEA also showed positive associations for peroxisome, bile acid metabolism and fatty acid metabolism. More HM-1 tumors showed negative enrichment of these three hallmark gene sets and related metabolites than HM-2 tumors (Fig. 6F). We observed a similar enrichment pattern (the sign of NES) for bile acid metabolism in nine of 11, i.e., ∼ 82% (considering only proteome data) or in 12 of 14, i.e., ∼ 86% (considering both proteome and transcriptome data) cases (Fig. 6F and 6G). Similar patterns for peroxisome and fatty acid metabolism, particularly in the HM-1 cases, were also observed (Fig. S6a). On the other hand, the negative enrichment of proteome-based xenobiotic metabolism gene set was associated with mostly positive enrichment of metabolome-based xenobiotic metabolites (primarily xenobiotics) in both HM-1 and HM-2 tumors (Fig. 6F, Fig. S6a). Thus, the loss of liver functions in bile acid, fatty acid, and xenobiotic metabolism revealed by transcriptome and proteome data was validated by decreased bile acids and fatty acids and increased xenobiotics in STX Hispanic HCC tumors.

While fatty acid metabolism was lower in the tumors compared to non-tumor tissues, as shown by the overall GSEA (Fig. 6E) and in more HM-1 tumors than in HM-2 tumors (Fig. S6a), we found that most of the very-long-chain fatty acids (VLCFAs) with chain-length of ≥ 20 carbons were relatively more accumulated in the HM-1 tumors, with the levels of many VLCFAs considerably higher in HM-1 cases than in HM-2 cases (Fig. S6b, top). This difference was significant for adrenate (or docosatetraenoate, 22:4) (p=0.029) and eicosapentaenoate (20:5) (p=0.043) (both are polyunsaturated fatty acids). Overall, both monounsaturated and polyunsaturated fatty acids were at significantly (Wilcoxon test, p<0.05) higher levels in HM-1 tumors but not in HM-2 tumors (Fig. S6b, bottom). Thus, there appears to be a relatively higher accumulation of unsaturated VLCFAs in the HM-1 tumors than in adjacent non-tumor tissues from the STX-Hispanic patients.

### Significant reduction of serum lipids in HCC patients

The significant negative enrichments of adipogenesis [in 14 of 22 (∼64%) STX-Hispanic HCC and 30 of 50 (=60%) TCGA-LIHC] and fatty acid metabolism [in 14 of 22 (∼64%) STX-Hispanic HCC and 41 of 50 (=82%) TCGA-LIHC] in the majority of HCC tumors revealed by ssGSEA of both transcriptome and proteome data (Fig. 4A) led us to compare serum lipid profiles between STX Hispanic and STX non-Hispanic individuals with or without HCC (Fig. 1B). We collected serum samples from 20 HCC patients (10 Hispanic, 10 non-Hispanic) and 20 non-HCC individuals (10 Hispanic, 10 non-Hispanic). PCA analysis based on 289 serum lipids separated HCC patients from non-HCC individuals except for two non-HCC cases (TCB_02056 and TCB_16096) (Fig. 7A). A retrospective patient clinical data review found evidence of cirrhosis and/or steatosis for the two abnormal cases (Fig. 1B). They were considered outliers and not included in subsequent analyses. We found no significant difference in age and BMI among the four groups separated based on ethnicity and HCC status (Fig. S7a).

**Figure 7:**
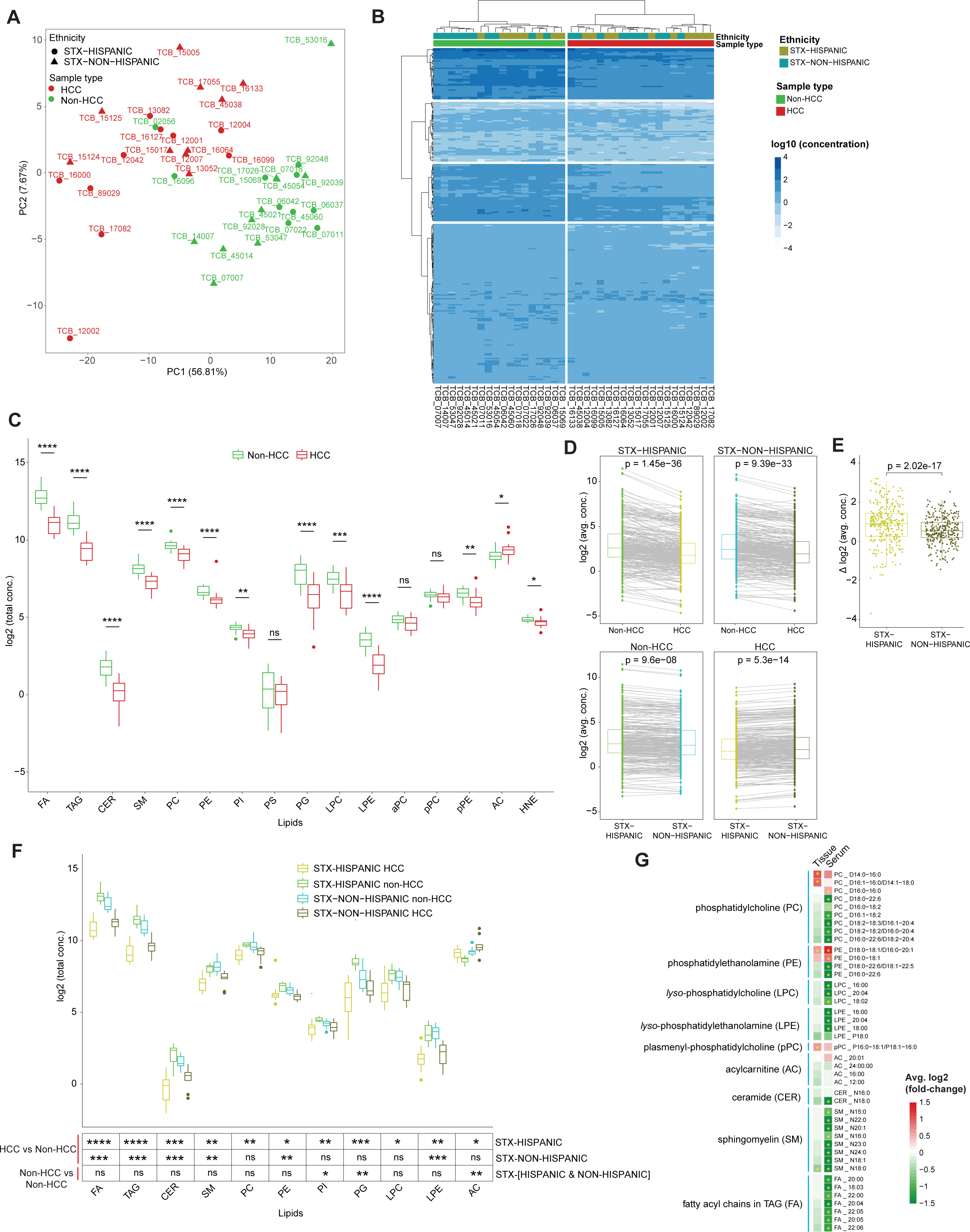
Dysregulated lipids in the serum samples of HCC patients from different ethnic populations. (A) PCA of serum lipid profiles from HCC and non-HCC individuals from Hispanic and non-Hispanic donors. Lipid concentrations were log10 transformed before analysis. (B) Independent of ethnicity, HCC (n=20) shows different lipid profiles than non-HCC individuals (n=18). (C) Comparison of the total concentration of 16 profiled lipid types between HCC and non-HCC. (D) Trends of lipid concentrations between HCC and non-HCC samples, and between Hispanic and non-Hispanic samples. Each line connects the same lipids. The dot represents the average of the subjects in the group. (E) Boxplot representation of Hispanics vs. non-Hispanics for differences in log2 transformed average concentration [Δ log2 (avg. conc.)] of 289 lipids between non-HCC and HCC. (F) Distribution of total lipid concentrations for individual lipid types across four datasets. We applied two-sided Wilcoxon rank-sum tests to determine statistical significance for all comparisons in C and E. The p-values in D were calculated using paired sample t-tests. (G) Selected lipids showing a similar direction of dysregulation in HCC tumor vs. adjacent non-tumor tissues and HCC vs. non-HCC serum samples in STX-Hispanics. Abbreviations of serum lipids: FA, fatty acyl chains in TAG; TAG, triacylglycerol; CER, ceramide; SM, sphingomyelin; PC, phosphatidylcholine; PE, phosphatidylethanolamine; PI, phosphatidylinositol; PS, phosphatidylserine; PG, phosphatidylglycerol; LPC, *lyso*-phosphatidylcholine; LPE, *lyso*-phosphatidylethanolamine; aPC, plasmanyl-phosphatidylcholine; pPC, plasmenyl-phosphatidylcholine; pPE, ethanolamine plasmalogens; AC, acylcarnitine; HNE, 4−hydroxy nonenal.

We found that HCC could be clearly distinguished from non-HCC individuals independent of patient ethnicity (Fig. 7B). Total lipid concentration of 16 lipid-types (Fig. S7b) was obtained by adding individual lipid concentrations within the same type (number of lipids within each lipid-types ranged between 2 and 100) for every sample. We examined the difference in the total lipid concentrations of the 16 lipid types between HCC and non-HCC cases. Seventy-five percent of the serum lipid levels were significantly lower while only acylcarnitines (AC) was significantly higher in HCC patients than in non-HCC individuals (Fig. 7C).

Comparing the mean level of each lipid between groups based on ethnicity and HCC status, we observed that HCC patients had a significantly lower abundance of lipids than non-HCC individuals regardless of ethnicity (Fig. 7D). However, the majority of lipids from HCC patients were more depleted in STX-Hispanics than in STX-non-Hispanics (paired sample t-test, p=2.02e-17) (Fig. 7E). Among the individuals without HCC, STX-Hispanics had a significantly higher abundance of serum lipids than STX-non-Hispanics (Fig. 7D lower left). In contrast, among the patients with HCC, STX-Hispanics had a significantly lower abundance of serum lipids than STX-non-Hispanics (Fig. 7D, lower right).

The difference between the two ethnic groups is further illustrated in Fig. 7F, which shows significantly lower levels of 10 serum lipid types and higher AC in HCC than non-HCC STX-Hispanics. In contrast, only five serum lipid types were significantly lower in HCC than non-HCC STX-non-Hispanics. Finally, forty-two lipids belonging to nine lipid types showed a similar direction of dysregulation, mainly reduction, in tumor compared with paired non-tumor (tissue metabolomic) and in serum collected from HCC patients compared with serum from non-HCC subjects in STX-Hispanics (Fig. 7G). Taken together, STX-Hispanics exhibited more reductions of their serum lipids in HCC subjects than STX-non-Hispanics, likely due to more impairment of their liver function.

## Discussion

Our cohort comprised 31 self-reported Hispanic patients from South Texas. We confirmed the genetic ancestry of every HCC patient in our cohort whose WES data was available. We found significantly more *AXIN2* mutations in our STX-Hispanic cohort than in TCGA-LIHC non-Hispanic Whites and other non-US-based ICGC cohorts, including LICA−FR, LINC−JP, and LIRI−JP. The loss of function mutation of *AXIN2* is significant as it is a downstream target and negative feedback regulator of Wnt/β-catenin pathway [67]. The frequency of gain-of-function mutations in *CTNNB1* was also higher in STX-Hispanic than other ethnic groups in TCGA-LIHC by 7-15%, except the African American group. Genomic alterations revealed Wnt/β-catenin as the most recurrently altered signaling pathway in STX-Hispanic HCC. Wnt/β-catenin pathway, of which *CTNNB1*, *AXIN1*, and *AXIN2* are key components, has critical roles in various steps of hepatocarcinogenesis [68]. These observations suggest that constitutive activation of Wnt/β-catenin pathway due to *AXIN2* and *CTNNB1* mutation may play a dominant role in HCC tumorigenesis among STX Hispanics [69]. The frequency of *TERT* promoter mutations, another known driver of hepatocarcinogenesis, was higher in STX-Hispanic HCC cohort than the HCC from other ethnicities. The difference was significant in comparison to TCGA-LIHC Asian and non-Hispanic White cohorts, and a non-US-based Asian population. mTOR signaling is known to be upregulated in HCC [70]. *MTOR* mutations were detected and validated in the STX-Hispanic HCC cohort at a higher frequency than in other ethnic groups in the TCGA-LIHC cohort, except the African-American group. The mutation frequency of *TP53* was lower in the STX-Hispanic HCC tumors than those of different ethnicities.

We identified ageing related signatures SBS1 and SBS5 [33,71], and recurrent signatures related to aflatoxin exposure (SBS24) and tobacco chewing (SBS29)[33,71]. Unlike TCGA-LIHC [33], we observed APOBEC (SBS2), homologous recombination deficiency (SBS3), UV exposure (SBS7b), and other unknown (SBS39) signatures in our STX-Hispanic cohort. Thus, our results show ubiquitous as well as unique mutation signatures in STX-Hispanic HCC. Studies with larger cohorts are needed to validate these mutation signatures in Hispanic HCC.

Unsupervised clustering of patients, using the enrichment scores of 50 hallmark gene sets from either transcriptome-or proteome-based ssGSEA, divided the same STX-Hispanic HCC cases into two groups, indicating the utility of genome-wide proteomic data for clustering diseases with different properties. The same clustering pattern of our Hispanic cohort was also observed in the TCGA-LIHC cohort. Regardless of ethnicity, the majority of tumors from the two clusters (i.e., HM-1 and HM-2) were positively or negatively enriched, respectively, for immune- and angiogenesis-related gene sets. The significantly better survival of HM-1 patients was likely due to elevated host immune killing of tumor cells and better response to anti-angiogenesis therapy such as sorafenib.

The significant enrichment of anti-tumor immune cells including NK cells, CD4+ and CD8+ T-cells in HM-1 tumors suggests that HM-1 tumors were likely suppressed by host immune surveillance [72], which might have contributed to the better survival of the HM-1 patients. HM-1 tumors also have more exhausted (Tex) and regulatory (Tregs) T-cells than HM-2 tumors, which could attenuate the anti-tumor immunity imparted by the helper and cytotoxic T-cells. Our observation of elevated levels of many VLCFAs in HM-1 tumors in contrast to HM-2 tumors might explain a possible mechanism for exhaustion of effector T-cells in the HM-1 patients. Enriched free polyunsaturated fatty acids (PUFAs) in the tumor tissues is known to induce CD8+ T cell ferroptosis and loss of effector function [73]. Therefore, our observation of high PUFA in the HM-1 tumors is consistent with higher Tex cells in HM-1 tumors. In addition, most of the immune checkpoint receptors, including *PDCD1* (*PD-1*) and *CTLA4*, were upregulated in HM-1 and downregulated in HM-2 tumors in comparison to their adjacent non-tumor tissues in STX-Hispanic and TCGA-LIHC cohorts. Similar mRNA expression difference was also observed in their ligands, including *CD274* (*PD-L1*), *PDCD1LG2* (*PD-L2*), *CD80*, and *CD86*, between the two subtypes of tumors. These observations plus the positively correlated expression between the receptor and ligand pairs, including *PD-1/PD-L1* or *PD-L2*, *CTLA4/CD80* or *CD86*, and *BTLA*/*HVEM*, suggest that HM-1 tumors should be more responsive than HM-2 tumors to immune checkpoint inhibition therapy.

BTLA/HVEM may be novel targets for treating HM-1 tumors with checkpoint inhibition [74].

Our observation is consistent with the reports that Wnt/β-catenin pathway activation is associated with resistance to immune checkpoint inhibition therapy [75], as our data showed that HM-2 tumors are more frequently associated with mutational Wnt pathway activation, particularly in the STX-Hispanic cohort. Studies have shown that tumors with high TMB (usually TMB>10 mutations/Mb) respond better to PD-1/PD-L1 inhibitors [76]. However, we found that TMB in HM-1 and HM-2 tumors of both cohorts was lower than 10 mutations/Mb. Thus, while immune checkpoint inhibitors (ICIs) have shown their efficacy and are part of the frontline therapy for advanced HCC, Hispanic patients with HCC may be less responsive to ICIs than HCC patients from other ethnic groups given the high frequency of somatic mutations of the Wnt/β-catenin pathway components and the low TMB in the Hispanic HCC patients.

The analysis of our transcriptome and proteome data with ssGSEA indicated a general impairment of liver function in HCC tumors. This was also the case in the TCGA-LIHC. When metabolomic profiles of tumors were compared to adjacent non-tumor tissues of STX-Hispanics, we identified 11 significantly enriched pathways, including vitamin B6; nicotinate and nicotinamide; glutamate metabolisms; TCA cycle; and glycolysis and gluconeogenesis. Loss of B vitamins, including B6 (pyridoxine) and B3 (nicotinate or niacin), were previously reported to be associated with hepatocarcinogenesis [77,78]. Metabolites involved in the nicotinate and nicotinamide metabolism process, TCA cycle, and glycolysis and gluconeogenesis were predominantly found at a lower levels in HCC tumors than in adjacent non-tumor tissues, which was associated with significant negative enrichments of these pathways based on the GSEA results from the proteome data of our Hispanic HCC. Significantly lower abundances of phosphoenolpyruvate (PEP), and the rate-limiting hepatic gluconeogenic enzyme PCK1, was seen in tumors than in adjacent non-tumor tissue samples of our HCC cohort. Reduction of PCK1 in our HCC tumors was significantly positively correlated with PEP levels. PCK1 plays a key role in controlling gluconeogenesis and TCA cycle maintenance by regulating TCA cataplerosis [79]. Depletion of PCK1 was linked with hyper-O-GlcNAcylation leading to liver oncogenesis [80]. The metabolism of the glutamate formed in the liver can be via either glutamate dehydrogenase or transamination and subsequent flux to urea synthesis and gluconeogenesis [81].

We observed significant negative enrichments of bile acid and fatty acid metabolisms in both proteome and metabolome profiles of STX-Hispanic HCC. A recent study has shown that aberrant bile acid metabolism facilitates HCC development by preventing natural killer T (NKT) cell recruitment and increasing M2-like tumor-associated macrophage polarization [82]. Dysregulation of fatty acid metabolism, an important metabolic rewiring phenomenon in tumor cells and immunocytes, is known to be involved in HCC development and progression [19]. We observed significant reduction of carnitine palmitoyltransferase 2 (CPT2), which converts acylcarnitine to acyl-CoA, by 1.7-fold both at the transcript and protein levels of STX-Hispanic HCC tumors in comparison with adjacent non-tumor tissue samples. There was a 2.4-fold reduction in the expression of SLC25A20 (also known as CACT), a mitochondrial-membrane carrier protein that helps in the translocation of acylcarnitine to the mitochondria. Relatively low abundance of CPT2 in tumor tissues causes HCC cells to escape from lipotoxicity by suppressing fatty acid β-oxidation pathway and accumulation of acylcarnitine (AC) in HCC serum samples [19]. Relatively less abundant CACT and CPT2, resulting in reduced acyl-CoA production from acylcarnitine, and marked accumulation of the latter, was identified in high-fat diet-induced HCC [83]. We also observed significantly elevated acylcarnitine in the serum profiles of Hispanic HCC. Thus, the revelation of these aberrant metabolites and pathways may lead to identifying potential targets for diagnosis and treating HCC, particularly in Hispanics.

Our serum lipid profiling showed significantly lower levels of most serum lipid types in HCC patients compared to non-HCC individuals, more so in Hispanic than in non-Hispanic patients. Acylcarnitine was the only lipid type whose level was significantly higher in HCC patients. This could be due to the substantial reduction of CPT2 in HCC tumors as discussed above. Similar to our observations, levels of serum triacylglycerols, phosphatidylcholines, and *lyso*-phosphatidylcholines were found to be lower in HBV-related HCC than in healthy controls [84]. HCC patients were found to have reduced levels of phosphatidylinositols, phosphatidylglycerols, and non-esterified fatty acids relative to healthy normal or patients with chronic liver disease [18]. These six lipid classes were prominently more reduced in serum samples of Hispanic than non-Hispanic HCC patients. Our results suggest a higher degree of dysregulation of serum lipid profiles of STX-Hispanic HCC compared with STX-non-Hispanic HCC patients.

## Conclusions

Our integrative multi-omics analyses of Hispanic HCC revealed some unique molecular features of Hispanic HCC, including a high mutation frequency of the *TERT* promoter and predominant activation of the Wnt/β-catenin signaling pathway due to the high mutation frequency of *AXIN2* and *CTNNB1,* with implications for therapy responsiveness. Our study also revealed significant reduction of most serum lipids in HCC patients, which was concomitant with dysregulation of many key hepatic enzymes involved in lipid metabolism, and with significant negative enrichment of adipogenesis and fatty acid metabolisms in the HCC tumors. Our study also revealed some common features between Hispanic and non-Hispanic HCC, such as significant aberration of the p53 pathway, activation of cell cycle pathways with positive enrichment of cell cycle-related gene sets, and significant negative enrichment of gene sets related to liver metabolic functions. Our application of ssGSEA and unsupervised clustering of patients using the enriched hallmark gene sets add prognostic value by identifying two subtypes of HCC with distinctive opposite enrichment between immune-related and liver-function related gene sets. The clustering may be of value for the prediction of response to immune checkpoint and angiogenesis inhibition therapy.

The small sample size is a main limitation of our study. Nevertheless, because our study involved the comparisons between paired tumor and adjacent non-tumor tissues for the multiple omics analyses, we did observe many statistically significant changes during the development and progression of the STX-Hispanic HCC with some important molecular profiles different from those of other ethnic HCCs. Further studies on a large Hispanic HCC cohort will help to validate our findings. Since Hispanic patients are notably underrepresented in all cancer genomic cohorts that are available, our multiple omics data sets on Hispanic HCC provide a unique resource for future research on this population.

## Data availability

The aligned BAM files from tumor-nontumor paired samples of 27 STX-Hispanic HCC patients (discovery cohort) were deposited to the European Genome-phenome Archive (EGA) under study accession id <EGAS00001007431> (for use after the dataset is made public). The aligned BAM files for RNA-Seq data from the 13 tumor−nontumor paired samples are available from the GEO database with accession # GSE233422. The raw mass spectrometry based proteomic data from 15 tumor−nontumor paired samples can be accessed from MassIVE (http://massive.ucsd.edu) with accession id <PXD043687> (for use after the dataset is made public). Other analyzed MS-based data files are available from the link https://doi.org/10.7303/syn51591313.

## Authors’ Contributions

Conceptualization of the study: DD, XW, YC, SZ, FGC, LZS. Supervision: SZ, LZS. Funding acquisition and project administration: FGC, LZS. Analysis: DD, XW, YCC, MC, JEM, YC. Investigation: DD, XW, YCC, HB, ZL, YZ, HIHC, CRZT, XG, MC, GAH, YC, SZ, FGS, LZS. Methodology: DD, XW, YCC, HB, FES, ZL, STW, XH, XG, JEM, YC, SZ, LZS. Sample collection: HB, JEL, CRZT, GAH, FGC. Sample examination and evaluation: FES. Genome and transcriptome data acquisition: ZL. Proteome data acquisition: STW. Lipidome data acquisition: XH. Writing of original draft: DD, LZS. Review and editing of draft: DD, XW, YCC, HB, FES, JEL, ZL, STW, XH, JEM, YC, SZ, FGC, LZS. All authors read and approved the final manuscript.

## Acknowledgements

This work was supported by funding from the Clayton Foundation for Research to FGC and L-Z S, by NIH cancer center support grant P30 CA054174 to the Shared Resources of Mays Cancer Center’s Next Generation Sequencing and Mass Spectrometry. Next Generation Sequencing Shared Resource is also supported by NIH Shared Instrument grant S10 OD030311 and CPRIT Core Facility Award (RP220662), and R50 CA265339-01A1 to Z. Lai. Mass spectrometry analyses were also supported in part by the University of Texas System Proteomics Core Network for purchase of the Orbitrap Fusion Lumos mass spectrometer, with expert technical assistance of Sammy Pardo and Dana Molleur. The Functional Lipidomics Core at the University of Texas Health Science Center at San Antonio was partially supported by P30 AG013319 and P30 AG044271. CRZ was supported by NIH T32CA148724 and F32CA228435 training grants. We gratefully acknowledge all the patients who participated in the study, our lab members (Junhua Yang and Araceli S Huerta for technical support), radiologists in the Department of Radiology for patient sample collection, and transplant center research coordinators, Sarah Schwartz, Michelle Brady, Rani Meenakshi, Stephanie Hargis, Stephanie Milroy, and Jillian Woodworth, for patient sample collection and deidentified demographic data. The TCGA-LIHC datasets used for the analyses described in this manuscript were obtained from dbGap at https://www.ncbi.nlm.nih.gov/gap/ through dbGap accession phs000178.v10.p8 under the South Texas Liver Cancer Study (project ID: 13485).

## References

1. Sung H, Ferlay J, Siegel RL, Laversanne M, Soerjomataram I, Jemal A, et al. Global Cancer Statistics 2020: GLOBOCAN Estimates of Incidence and Mortality Worldwide for 36 Cancers in 185 Countries. CA Cancer J Clin. 2021;71:209–49.

2. Siegel RL, Miller KD, Wagle NS, Jemal A. Cancer statistics, 2023. CA Cancer J Clin. 2023;73:17–48.

3. Asafo-Agyei KO, Samant H. Hepatocellular Carcinoma. 2022.

4. El-Serag HB, Sardell R, Thrift AP, Kanwal F, Miller P. Texas Has the Highest Hepatocellular Carcinoma Incidence Rates in the USA. Dig Dis Sci. 2021;66:912–6.

5. Thrift AP, Liu KS, Raza SA, El-Serag HB. Recent Decline in the Incidence of Hepatocellular Carcinoma in the United States. Clinical Gastroenterology and Hepatology. 2022;

6. Alemán JO, Almandoz JP, Frias JP, Galindo RJ. Obesity among Latinx people in the United States: A review. Obesity (Silver Spring). 2023;31:329–37.

7. Marengo A, Rosso C, Bugianesi E. Liver Cancer: Connections with Obesity, Fatty Liver, and Cirrhosis. Annu Rev Med. 2016;67:103–17.

8. Caldwell SH, Crespo DM, Kang HS, Al-Osaimi AMS. Obesity and hepatocellular carcinoma. Gastroenterology. 2004;127:S97–103.

9. Gupta A, Das A, Majumder K, Arora N, Mayo HG, Singh PP, et al. Obesity is Independently Associated With Increased Risk of Hepatocellular Cancer–related Mortality. Am J Clin Oncol. 2018;41:874–81.

10. Tan DSW, Mok TSK, Rebbeck TR. Cancer Genomics: Diversity and Disparity Across Ethnicity and Geography. J Clin Oncol. 2016;34:91–101.

11. Candia J, Bayarsaikhan E, Tandon M, Budhu A, Forgues M, Tovuu L-O, et al. The genomic landscape of Mongolian hepatocellular carcinoma. Nat Commun. 2020;11:4383.

12. Ally A, Balasundaram M, Carlsen R, Chuah E, Clarke A, Dhalla N, et al. Comprehensive and Integrative Genomic Characterization of Hepatocellular Carcinoma. Cell. 2017;169:1327–1341.e23.

13. Ng CKY, Dazert E, Boldanova T, Coto-Llerena M, Nuciforo S, Ercan C, et al. Integrative proteogenomic characterization of hepatocellular carcinoma across etiologies and stages. Nat Commun. 2022;13:2436.

14. Gao Q, Zhu H, Dong L, Shi W, Chen R, Song Z, et al. Integrated Proteogenomic Characterization of HBV-Related Hepatocellular Carcinoma. Cell. 2019;179:561–577.e22.

15. Zhuang W, Sun H, Zhang S, Zhou Y, Weng W, Wu B, et al. An immunogenomic signature for molecular classification in hepatocellular carcinoma. Mol Ther Nucleic Acids. 2021;25:105–15.

16. Montironi C, Castet F, Haber PK, Pinyol R, Torres-Martin M, Torrens L, et al. Inflamed and non-inflamed classes of HCC: a revised immunogenomic classification. Gut. 2023;72:129–40.

17. Liu J, Geng W, Sun H, Liu C, Huang F, Cao J, et al. Integrative metabolomic characterisation identifies altered portal vein serum metabolome contributing to human hepatocellular carcinoma. Gut. 2022;71:1203–13.

18. Ismail IT, Elfert A, Helal M, Salama I, El-Said H, Fiehn O. Remodeling Lipids in the Transition from Chronic Liver Disease to Hepatocellular Carcinoma. Cancers (Basel). 2020;13:88.

19. Hu B, Lin J, Yang X, Sang X. Aberrant lipid metabolism in hepatocellular carcinoma cells as well as immune microenvironment: A review. Cell Prolif. 2020;53.

20. 1000 Genomes Project Consortium, Auton A, Brooks LD, Durbin RM, Garrison EP, Kang HM, et al. A global reference for human genetic variation. Nature. 2015;526:68–74.

21. Wang LJ, Zhang CW, Su SC, Chen HIH, Chiu YC, Lai Z, et al. An ancestry informative marker panel design for individual ancestry estimation of Hispanic population using whole exome sequencing data. BMC Genomics. 2019;20.

22. Li H, Durbin R. Fast and accurate short read alignment with Burrows-Wheeler transform. Bioinformatics. 2009;25:1754–60.

23. Li H, Handsaker B, Wysoker A, Fennell T, Ruan J, Homer N, et al. The Sequence Alignment/Map format and SAMtools. Bioinformatics. 2009;25:2078–9.

24. McKenna A, Hanna M, Banks E, Sivachenko A, Cibulskis K, Kernytsky A, et al. The Genome Analysis Toolkit: a MapReduce framework for analyzing next-generation DNA sequencing data. Genome Res. 2010;20:1297–303.

25. Koboldt DC, Zhang Q, Larson DE, Shen D, McLellan MD, Lin L, et al. VarScan 2: somatic mutation and copy number alteration discovery in cancer by exome sequencing. Genome Res. 2012;22:568–76.

26. Cibulskis K, Lawrence MS, Carter SL, Sivachenko A, Jaffe D, Sougnez C, et al. Sensitive detection of somatic point mutations in impure and heterogeneous cancer samples. Nat Biotechnol. 2013;31:213–9.

27. Wang K, Li M, Hakonarson H. ANNOVAR: functional annotation of genetic variants from high-throughput sequencing data. Nucleic Acids Res. 2010;38:e164–e164.

28. Lawrence MS, Stojanov P, Polak P, Kryukov G v., Cibulskis K, Sivachenko A, et al. Mutational heterogeneity in cancer and the search for new cancer-associated genes. Nature. 2013;499:214–8.

29. Kennedy SR, Schmitt MW, Fox EJ, Kohrn BF, Salk JJ, Ahn EH, et al. Detecting ultralow-frequency mutations by Duplex Sequencing. Nat Protoc. 2014;9:2586–606.

30. ICGC/TCGA Pan-Cancer Analysis of Whole Genomes Consortium. Pan-cancer analysis of whole genomes. Nature. 2020;578:82–93.

31. Nault JC, Mallet M, Pilati C, Calderaro J, Bioulac-Sage P, Laurent C, et al. High frequency of telomerase reverse-transcriptase promoter somatic mutations in hepatocellular carcinoma and preneoplastic lesions. Nat Commun. 2013;4:2218.

32. Blokzijl F, Janssen R, van Boxtel R, Cuppen E. MutationalPatterns: comprehensive genome-wide analysis of mutational processes. Genome Med. 2018;10:33.

33. Alexandrov LB, Kim J, Haradhvala NJ, Huang MN, Tian Ng AW, Wu Y, et al. The repertoire of mutational signatures in human cancer. Nature. 2020;578:94–101.

34. Mermel CH, Schumacher SE, Hill B, Meyerson ML, Beroukhim R, Getz G. GISTIC2.0 facilitates sensitive and confident localization of the targets of focal somatic copy-number alteration in human cancers. Genome Biol. 2011;12:R41.

35. Sanchez-Vega F, Mina M, Armenia J, Chatila WK, Luna A, La KC, et al. Oncogenic Signaling Pathways in The Cancer Genome Atlas. Cell. 2018;173:321–337.e10.

36. Chakravarty D, Gao J, Phillips SM, Kundra R, Zhang H, Wang J, et al. OncoKB: A Precision Oncology Knowledge Base. JCO Precis Oncol. 2017;2017.

37. Kim D, Pertea G, Trapnell C, Pimentel H, Kelley R, Salzberg SL. TopHat2: accurate alignment of transcriptomes in the presence of insertions, deletions and gene fusions. Genome Biol. 2013;14:R36.

38. Li B, Dewey CN. RSEM: accurate transcript quantification from RNA-Seq data with or without a reference genome. BMC Bioinformatics. 2011;12:323.

39. Love MI, Huber W, Anders S. Moderated estimation of fold change and dispersion for RNA-seq data with DESeq2. Genome Biol. 2014;15:550.

40. Robinson MD, McCarthy DJ, Smyth GK. edgeR: a Bioconductor package for differential expression analysis of digital gene expression data. Bioinformatics. 2010;26:139–40.

41. Torres-García W, Zheng S, Sivachenko A, Vegesna R, Wang Q, Yao R, et al. PRADA: pipeline for RNA sequencing data analysis. Bioinformatics. 2014;30:2224–6.

42. Haas BJ, Dobin A, Stransky N, Li B, Yang X, Tickle T, et al. STAR-Fusion: Fast and Accurate Fusion Transcript Detection from RNA-Seq. bioRxiv [Internet]. 2017;120295. Available from: http://biorxiv.org/content/early/2017/03/24/120295.abstract

43. Shen S, Park JW, Lu Z, Lin L, Henry MD, Wu YN, et al. rMATS: Robust and flexible detection of differential alternative splicing from replicate RNA-Seq data. Proceedings of the National Academy of Sciences. 2014;111.

44. Katz Y, Wang ET, Airoldi EM, Burge CB. Analysis and design of RNA sequencing experiments for identifying isoform regulation. Nat Methods. 2010;7:1009–15.

45. Colaprico A, Silva TC, Olsen C, Garofano L, Cava C, Garolini D, et al. TCGAbiolinks: an R/Bioconductor package for integrative analysis of TCGA data. Nucleic Acids Res. 2016;44:e71–e71.

46. Searle BC, Pino LK, Egertson JD, Ting YS, Lawrence RT, MacLean BX, et al. Chromatogram libraries improve peptide detection and quantification by data independent acquisition mass spectrometry. Nat Commun. 2018;9:5128.

47. Rosenberger G, Koh CC, Guo T, Röst HL, Kouvonen P, Collins BC, et al. A repository of assays to quantify 10,000 human proteins by SWATH-MS. Sci Data. 2014;1:140031.

48. Yu G, Wang L-G, Han Y, He Q-Y. clusterProfiler: an R package for comparing biological themes among gene clusters. OMICS. 2012;16:284–7.

49. Liberzon A, Birger C, Thorvaldsdóttir H, Ghandi M, Mesirov JP, Tamayo P. The Molecular Signatures Database Hallmark Gene Set Collection. Cell Syst. 2015;1:417–25.

50. Hoshida Y, Nijman SMB, Kobayashi M, Chan JA, Brunet J-P, Chiang DY, et al. Integrative Transcriptome Analysis Reveals Common Molecular Subclasses of Human Hepatocellular Carcinoma. Cancer Res. 2009;69:7385–92.

51. Yoshihara K, Shahmoradgoli M, Martínez E, Vegesna R, Kim H, Torres-Garcia W, et al. Inferring tumour purity and stromal and immune cell admixture from expression data. Nat Commun. 2013;4:2612.

52. Chen B, Khodadoust MS, Liu CL, Newman AM, Alizadeh AA. Profiling Tumor Infiltrating Immune Cells with CIBERSORT. Methods Mol Biol. 2018;1711:243–59.

53. Miao Y-R, Zhang Q, Lei Q, Luo M, Xie G-Y, Wang H, et al. ImmuCellAI: A Unique Method for Comprehensive T-Cell Subsets Abundance Prediction and its Application in Cancer Immunotherapy. Adv Sci (Weinh). 2020;7:1902880.

54. Ford L, Kennedy AD, Goodman KD, Pappan KL, Evans AM, Miller LAD, et al. Precision of a Clinical Metabolomics Profiling Platform for Use in the Identification of Inborn Errors of Metabolism. J Appl Lab Med. 2020;5:342–56.

55. Stekhoven DJ, Bühlmann P. MissForest--non-parametric missing value imputation for mixed-type data. Bioinformatics. 2012;28:112–8.

56. Jia P, Wang Q, Chen Q, Hutchinson KE, Pao W, Zhao Z. MSEA: detection and quantification of mutation hotspots through mutation set enrichment analysis. Genome Biol. 2014;15:489.

57. Pang Z, Chong J, Zhou G, de Lima Morais DA, Chang L, Barrette M, et al. MetaboAnalyst 5.0: narrowing the gap between raw spectra and functional insights. Nucleic Acids Res. 2021;49:W388–96.

58. Han X, Yang K, Gross RW. Multi-dimensional mass spectrometry-based shotgun lipidomics and novel strategies for lipidomic analyses. Mass Spectrom Rev. 2012;31:134–78.

59. Wang M, Han X. Multidimensional mass spectrometry-based shotgun lipidomics. Methods Mol Biol. 2014;1198:203–20.

60. Han X, Yang K, Gross RW. Microfluidics-based electrospray ionization enhances the intrasource separation of lipid classes and extends identification of individual molecular species through multi-dimensional mass spectrometry: development of an automated high-throughput platform f. Rapid Communications in Mass Spectrometry. 2008;22:2115–24.

61. Yang K, Cheng H, Gross RW, Han X. Automated Lipid Identification and Quantification by Multidimensional Mass Spectrometry-Based Shotgun Lipidomics. Anal Chem. 2009;81:4356–68.

62. Totoki Y, Tatsuno K, Covington KR, Ueda H, Creighton CJ, Kato M, et al. Trans-ancestry mutational landscape of hepatocellular carcinoma genomes. Nat Genet. 2014;46:1267–73.

63. Chang Y-S, Tu S-J, Chen H-D, Hsu M-H, Chen Y-C, Chao D-S, et al. Integrated genomic analyses of hepatocellular carcinoma. Hepatol Int. 2023;17:97–111.

64. Chiang DY, Villanueva A, Hoshida Y, Peix J, Newell P, Minguez B, et al. Focal Gains of VEGFA and Molecular Classification of Hepatocellular Carcinoma. Cancer Res. 2008;68:6779–88.

65. Gao J, Aksoy BA, Dogrusoz U, Dresdner G, Gross B, Sumer SO, et al. Integrative analysis of complex cancer genomics and clinical profiles using the cBioPortal. Sci Signal. 2013;6:pl1.

66. Bian X, Jiang H, Meng Y, Li Y, Fang J, Lu Z. Regulation of gene expression by glycolytic and gluconeogenic enzymes. Trends Cell Biol. 2022;32:786–99.

67. Bernkopf DB, Hadjihannas M V, Behrens J. Negative-feedback regulation of the Wnt pathway by conductin/axin2 involves insensitivity to upstream signalling. J Cell Sci. 2015;128:33–9.

68. Xu C, Xu Z, Zhang Y, Evert M, Calvisi DF, Chen X. β-Catenin signaling in hepatocellular carcinoma. J Clin Invest. 2022;132.

69. Carrot-Zhang J, Soca-Chafre G, Patterson N, Thorner AR, Nag A, Watson J, et al. Genetic Ancestry Contributes to Somatic Mutations in Lung Cancers from Admixed Latin American Populations. Cancer Discov. 2021;11:591–8.

70. Ferrín G, Guerrero M, Amado V, Rodríguez-Perálvarez M, De la Mata M. Activation of mTOR Signaling Pathway in Hepatocellular Carcinoma. Int J Mol Sci. 2020;21:1266.

71. Letouzé E, Shinde J, Renault V, Couchy G, Blanc J-F, Tubacher E, et al. Mutational signatures reveal the dynamic interplay of risk factors and cellular processes during liver tumorigenesis. Nat Commun. 2017;8:1315.

72. Lei X, Lei Y, Li J-K, Du W-X, Li R-G, Yang J, et al. Immune cells within the tumor microenvironment: Biological functions and roles in cancer immunotherapy. Cancer Lett. 2020;470:126–33.

73. Ma X, Xiao L, Liu L, Ye L, Su P, Bi E, et al. CD36-mediated ferroptosis dampens intratumoral CD8+ T cell effector function and impairs their antitumor ability. Cell Metab. 2021;33:1001–1012.e5.

74. Qin S, Xu L, Yi M, Yu S, Wu K, Luo S. Novel immune checkpoint targets: moving beyond PD-1 and CTLA-4. Mol Cancer. 2019;18:155.

75. Krutsenko Y, Singhi AD, Monga SP. β-Catenin Activation in Hepatocellular Cancer: Implications in Biology and Therapy. Cancers (Basel). 2021;13:1830.

76. Muhammed A, D’Alessio A, Enica A, Talbot T, Fulgenzi CAM, Nteliopoulos G, et al. Predictive biomarkers of response to immune checkpoint inhibitors in hepatocellular carcinoma. Expert Rev Mol Diagn. 2022;22:253–64.

77. Mei M, Liu D, Tang X, You Y, Peng B, He X, et al. Vitamin B6 Metabolic Pathway is Involved in the Pathogenesis of Liver Diseases via Multi-Omics Analysis. J Hepatocell Carcinoma. 2022;9:729–50.

78. Chedere A, Mishra M, Kulkarni O, Sriraman S, Chandra N. Personalized quantitative models of NAD metabolism in hepatocellular carcinoma identify a subgroup with poor prognosis. Front Oncol. 2022;12.

79. Liu M-X, Jin L, Sun S-J, Liu P, Feng X, Cheng Z-L, et al. Metabolic reprogramming by PCK1 promotes TCA cataplerosis, oxidative stress and apoptosis in liver cancer cells and suppresses hepatocellular carcinoma. Oncogene. 2018;37:1637–53.

80. Xiang J, Chen C, Liu R, Gou D, Chang L, Deng H, et al. Gluconeogenic enzyme PCK1 deficiency promotes CHK2 O-GlcNAcylation and hepatocellular carcinoma growth upon glucose deprivation. Journal of Clinical Investigation. 2021;131.

81. Watford M. Glutamine and Glutamate Metabolism across the Liver Sinusoid. J Nutr. 2000;130:983S–987S.

82. Xia JK, Tang N, Wu XY, Ren HZ. Deregulated bile acids may drive hepatocellular carcinoma metastasis by inducing an immunosuppressive microenvironment. Front Oncol. 2022;12:1033145.

83. Nakagawa H, Hayata Y, Kawamura S, Yamada T, Fujiwara N, Koike K. Lipid Metabolic Reprogramming in Hepatocellular Carcinoma. Cancers (Basel). 2018;10:447.

84. Chen S, Yin P, Zhao X, Xing W, Hu C, Zhou L, et al. Serum lipid profiling of patients with chronic hepatitis B, cirrhosis, and hepatocellular carcinoma by ultra fast LC/IT-TOF MS. Electrophoresis. 2013;n/a-n/a.

