## Supplementary Figures for "Integrative multi-omics characterization of hepatocellular carcinoma in Hispanic patients"

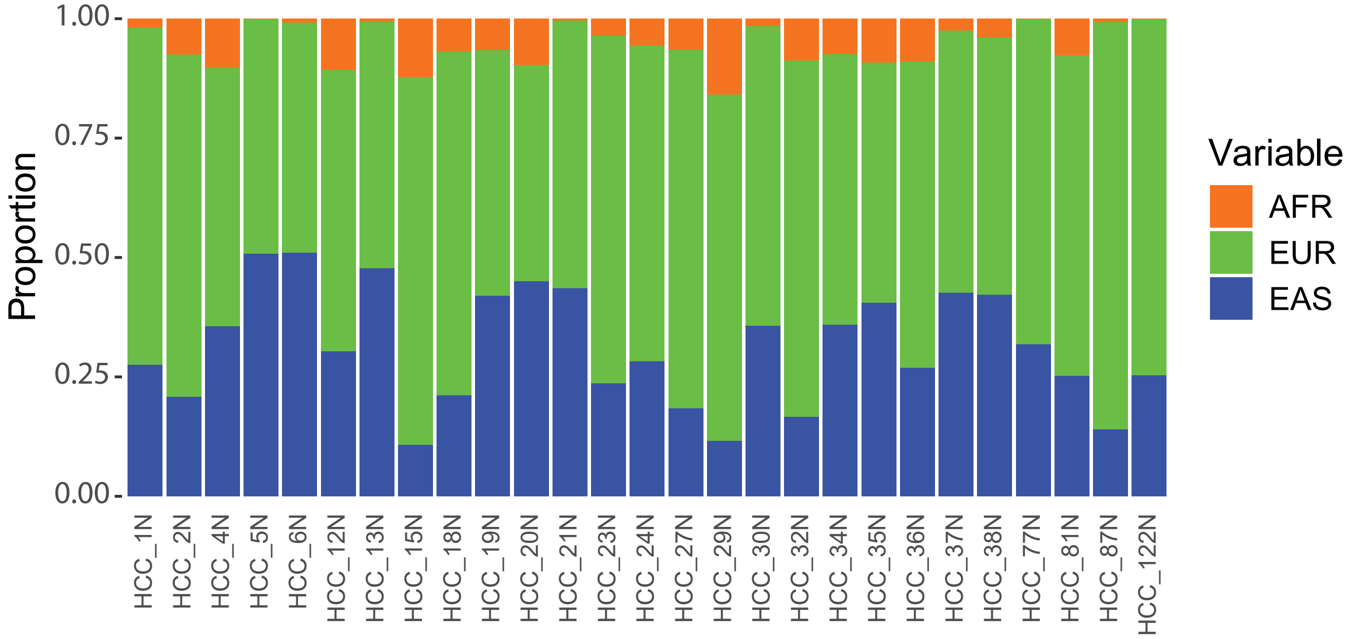


**Figure S1:** Plot show relative proportions of genetic ancestry coefficients derived from whole exome sequencing (WES) of non-tumor samples in our STX-Hispanic HCC cohort (n=27). Ancestry proportions are estimated from AFR: African, EUR: European, and EAS: East Asian. Patient ids are arranged in numerical order.

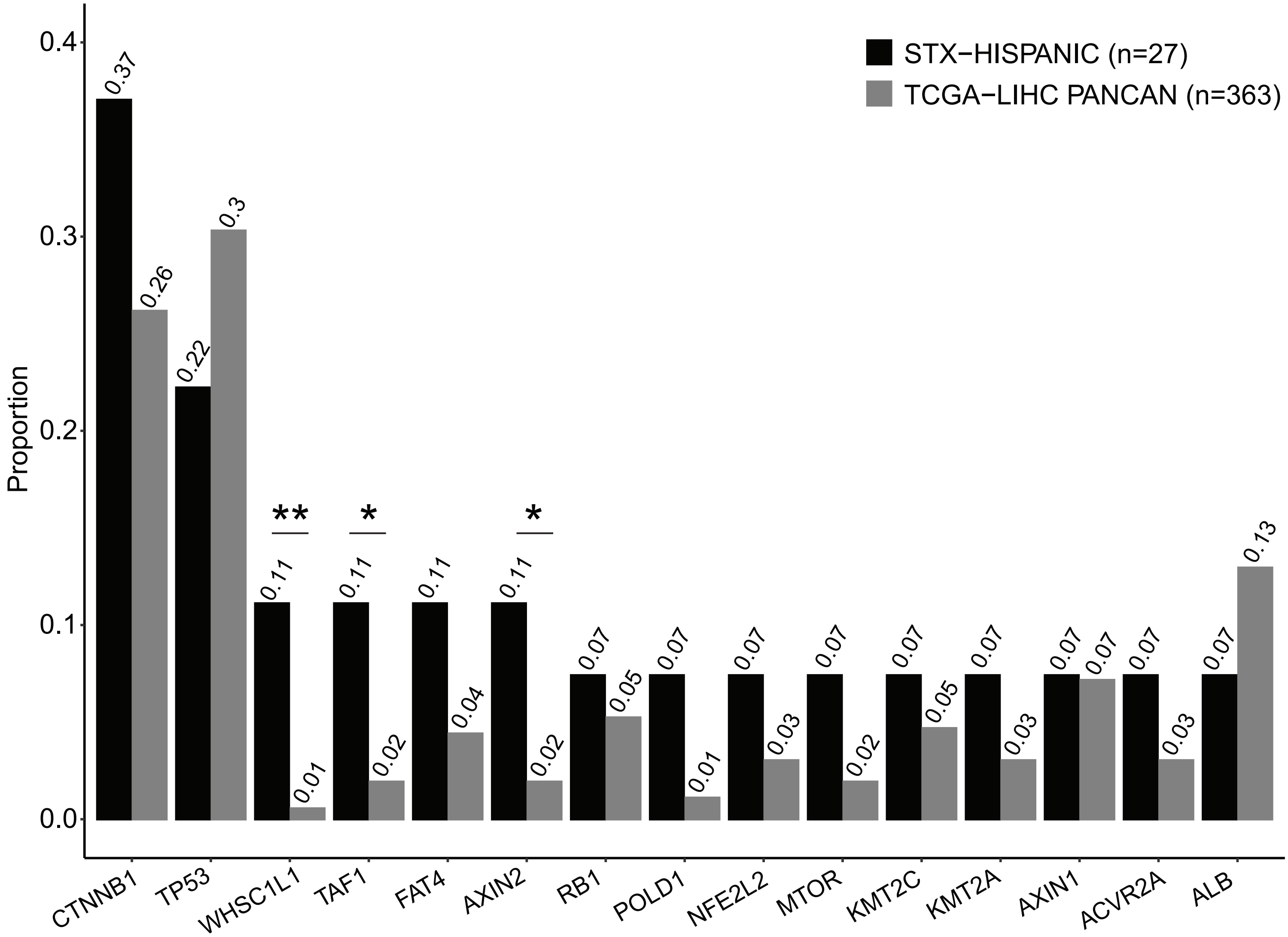


**Figure S2a:** Comparison of mutation frequencies of 15 recurrently mutated genes in HCC patients from STX-Hispanic and TCGA-LIHC cohorts reported in the pan-cancer studies (Fisher's exact test). Significant differences were represented by * (for 0.05 > p ≥ 0.01) and ** (for 0.01 > p ≥ 0.001).

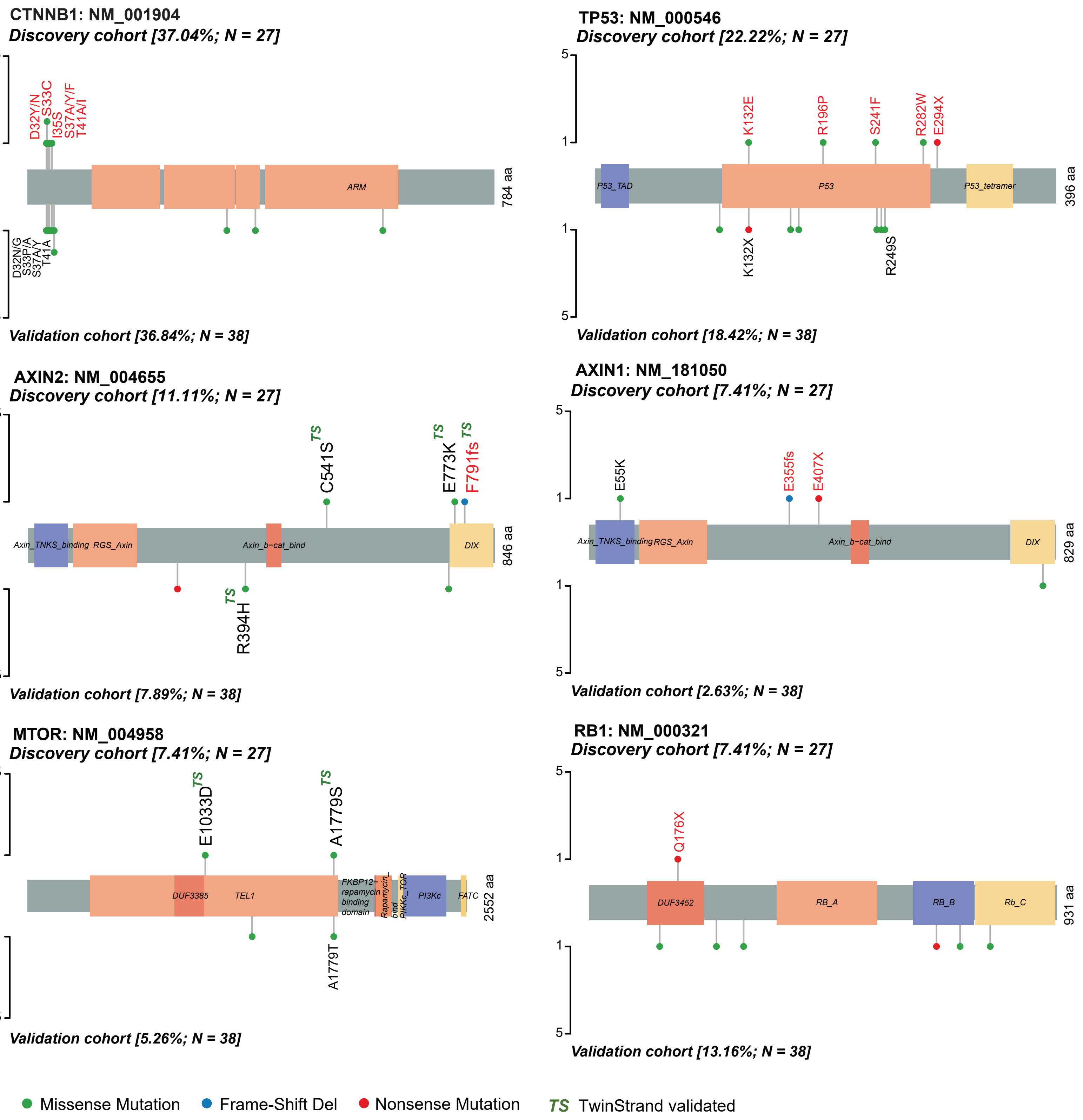


**Figure S2b:** Lollipop plots showing distributions of mutational hotspots in key genes in STX-Hispanic HCC. Variants identified in our 27 discovery and 38 validation HCC cohorts are indicated on the top and bottom of each plot, respectively. Amino acid (AA) changes found oncogenic/potentially oncogenic (OncoKB) are marked red. If a variant in the validation cohort was identified at the same locus as detected in discovery cohort patients, we have included corresponding AA changes in the plot. An aflatoxin-associated mutation in TP53 leading to R249S amino acid change seen in an HCC (validation cohort) patient has been marked. The superscript TS indicates AA changes validated using the DuplexSeq method.

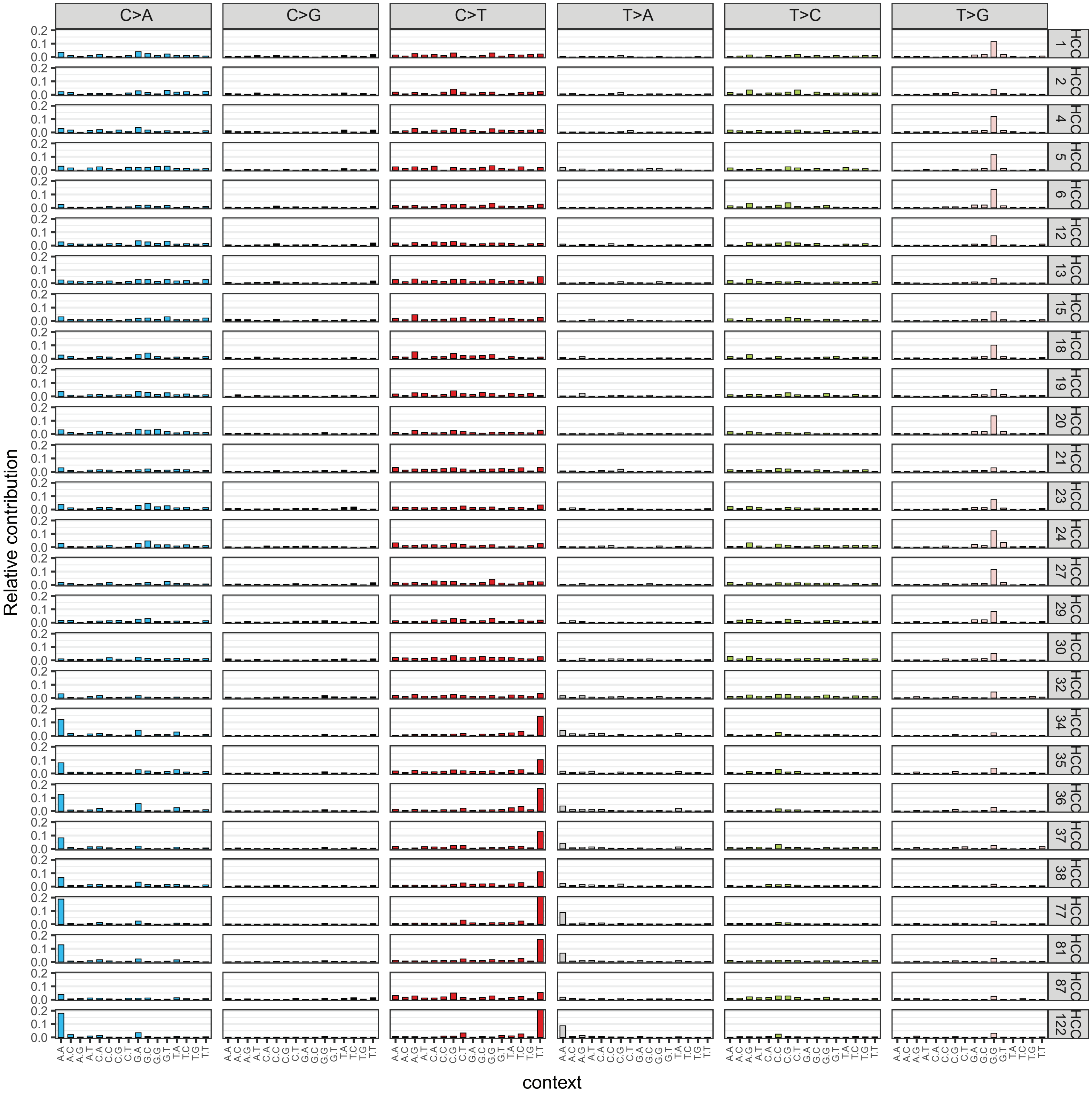


**Figure S2c:** Mutational spectra of 27 STX-Hispanic HCC (discovery cohort) across 96 mutation contexts. Patients are arranged in numerical order.

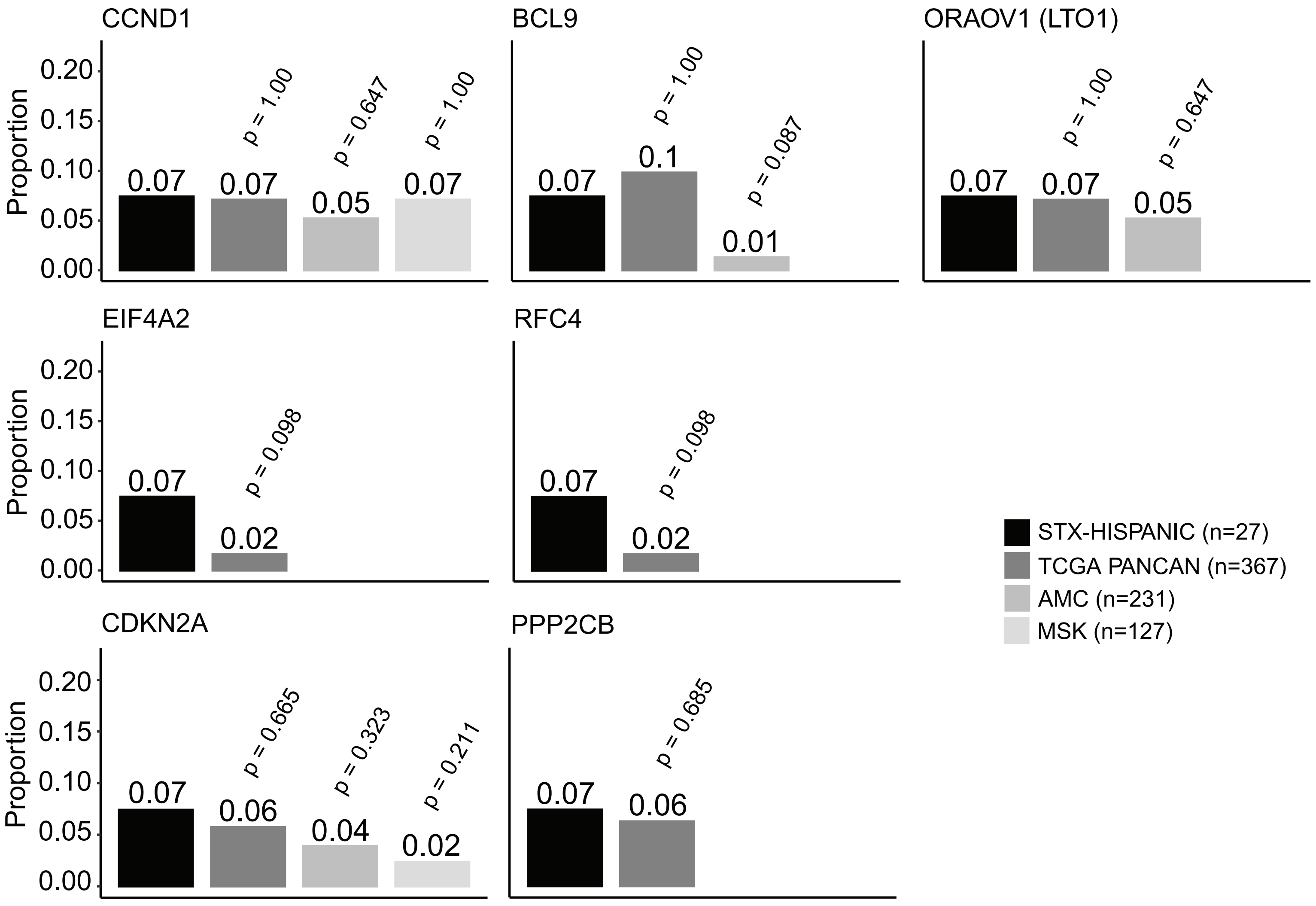


**Figure S2d:** Comparison of frequency of high-level copy number amplifications/deletions (GISTIC2.0 algorithm) between HCC tumors from STX-Hispanic and other study cohorts available in the cBio-Portal.

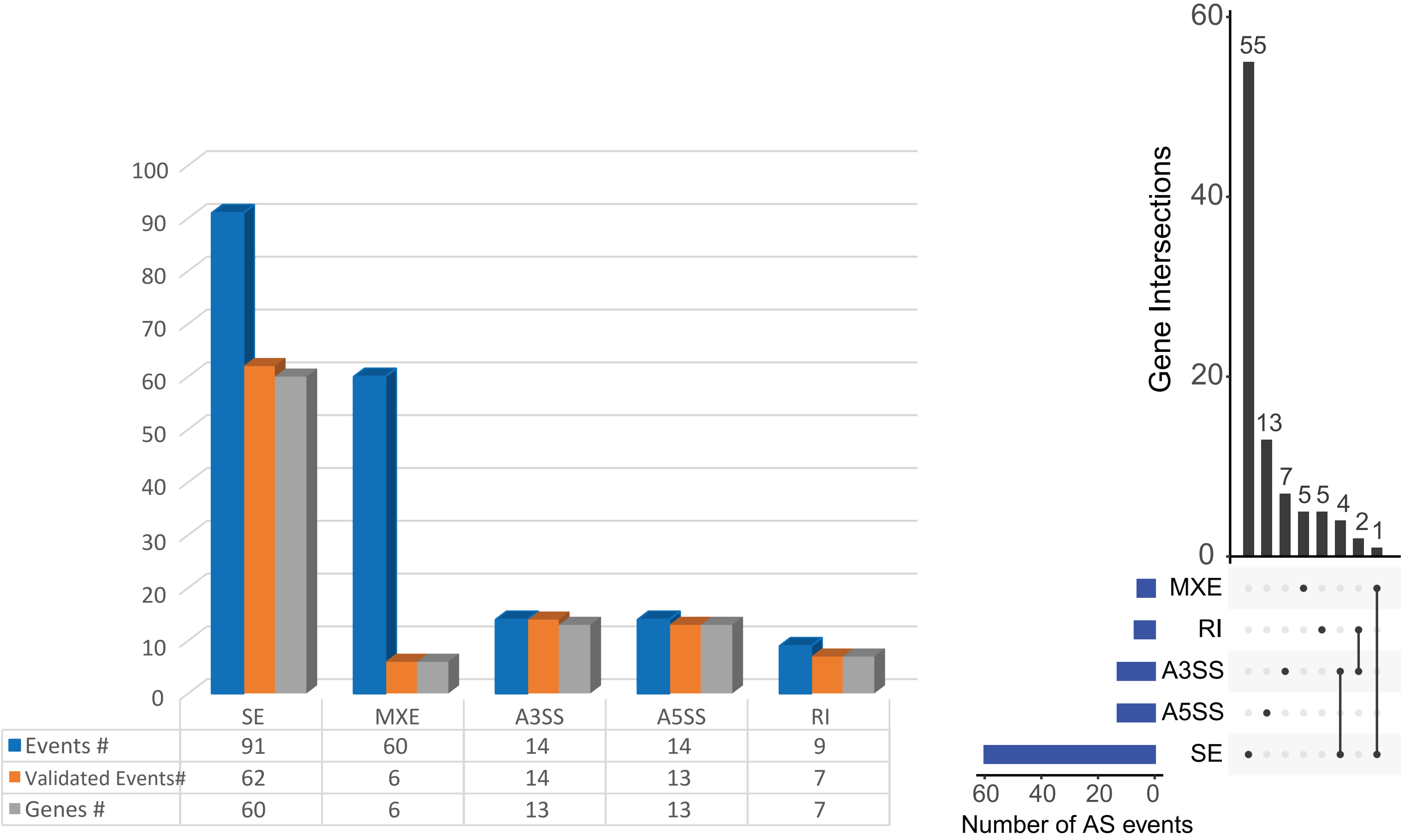


**Figure S3a:** Bar plot representation of the number of significant alternative splicing (AS) events identified using rMATS, AS events validated using MISO, and genes harboring the validated AS events (left panel). The upset plot on the right panel represents alternative splicing event patterns and genes identified and validated in our cohort of HCC tumors.

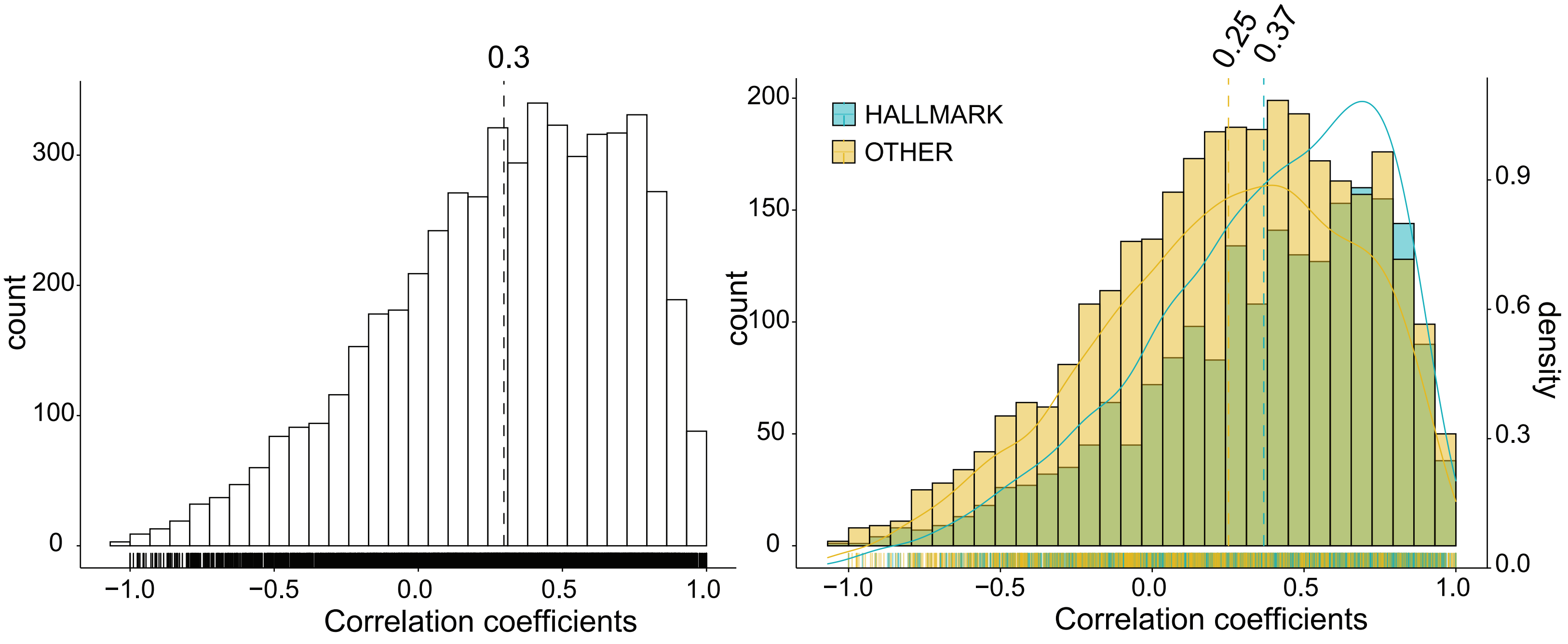


**Figure S3b:** Distribution of correlation coefficient between protein abundance and mRNA expression levels of 5460 gene-protein pairs from 6 STX-Hispanic HCC (left). On the right, the overlaying density plots show protein-mRNA pairwise correlation coefficients for hallmark (n=2123) and non-hallmark (n=3337) genes. The dashed lines indicate the mean correlation coefficient of all pairwise comparisons.

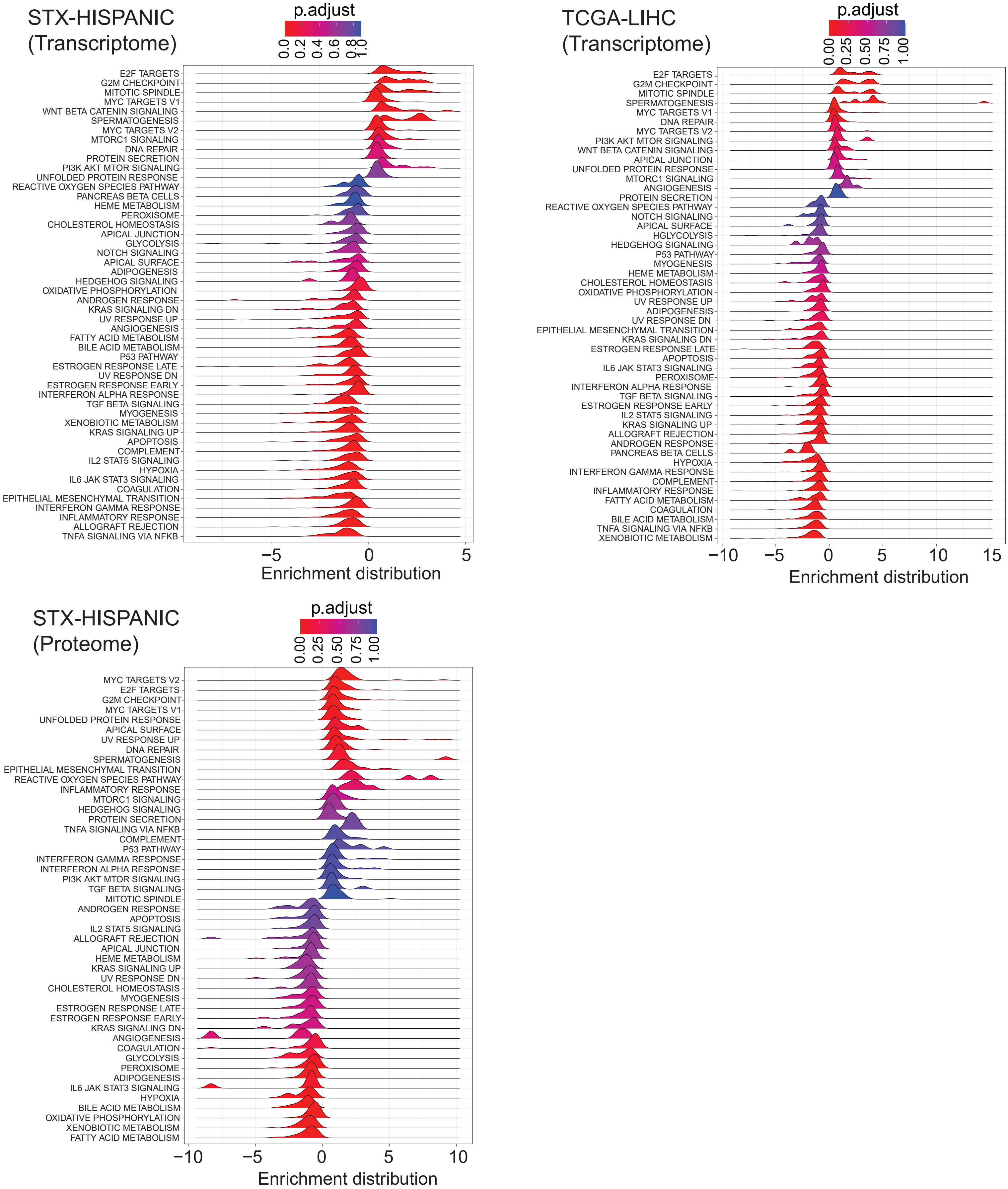


**Figure S3c:** Ridge plots depicting the distribution of enriched hallmark gene sets (n=50) in STX-Hispanic (top left) and TCGA (top right) HCC cohorts. The bottom left plot show enrichment distribution for 47 hallmark gene sets from gene set enrichment analysis (GSEA) on proteome data of STX-Hispanic HCC.

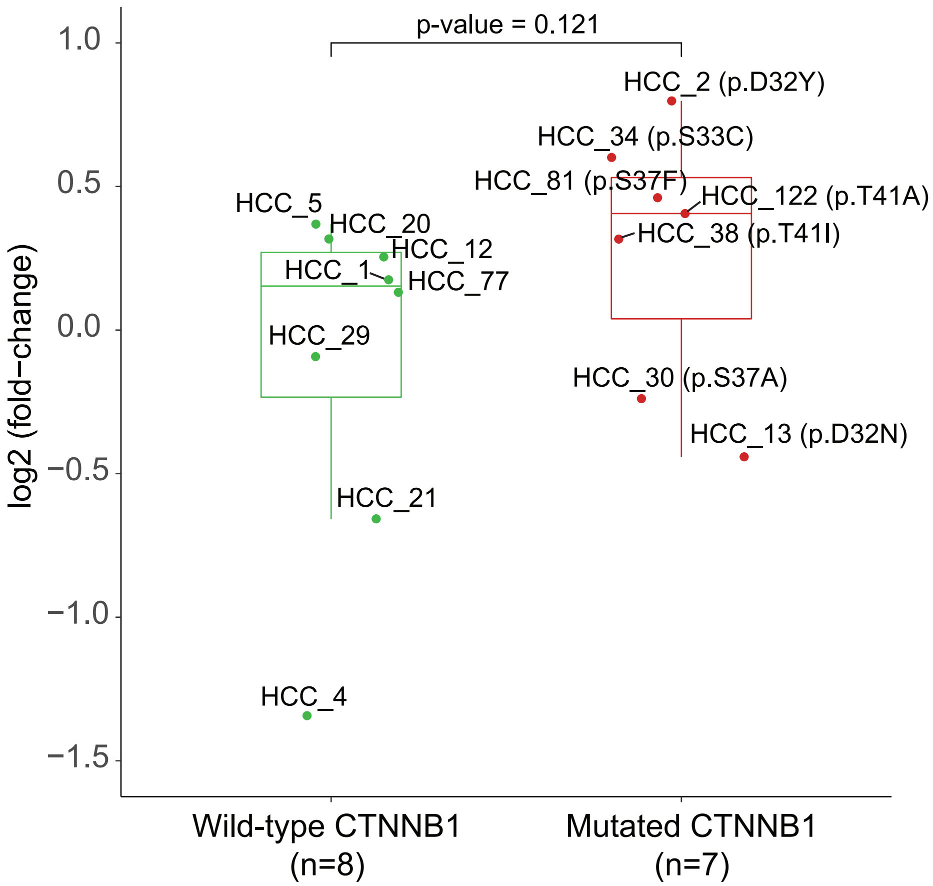


**Figure S3d:** Boxplot representation of log2 (fold-change) of mutated and wild-type CTNNB1 protein quantifications in HCC tumors compared to adjacent non-tumor tissue samples of STX-Hispanic HCC. The amino acid changes resulting from corresponding somatic mutations are indicated within parenthesis. The p-value was calculated using the Wilcoxon rank-sum test.

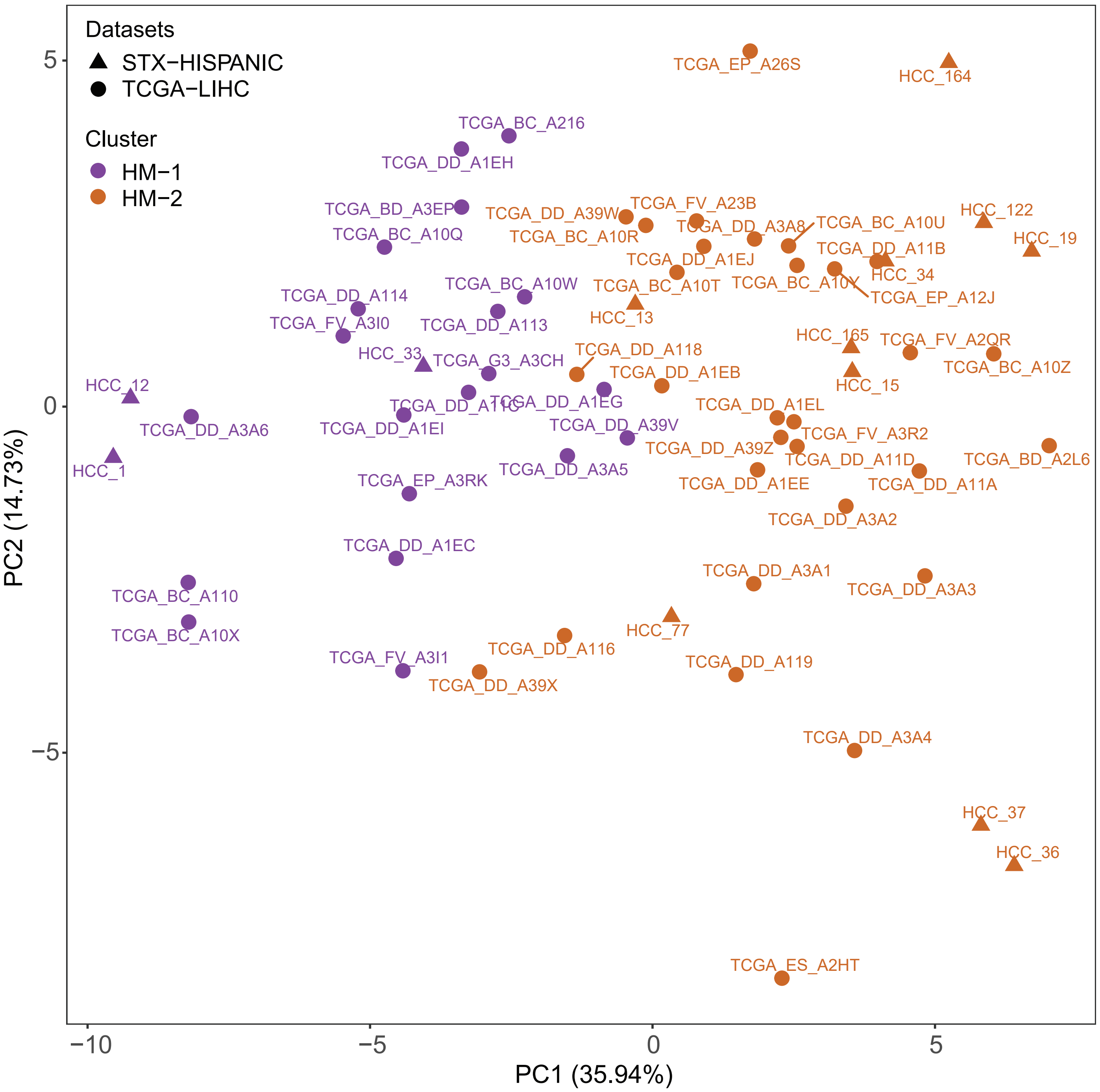


**Figure S4a:** Principal component analysis (PCA) plot to project HCC patients from HM-1 and HM-2 clusters onto the first two components. We used enrichment scores (NES) of 50 hallmark gene sets from 13 STX-Hispanic (triangle) and 50 TCGA (circle) HCC cohorts.

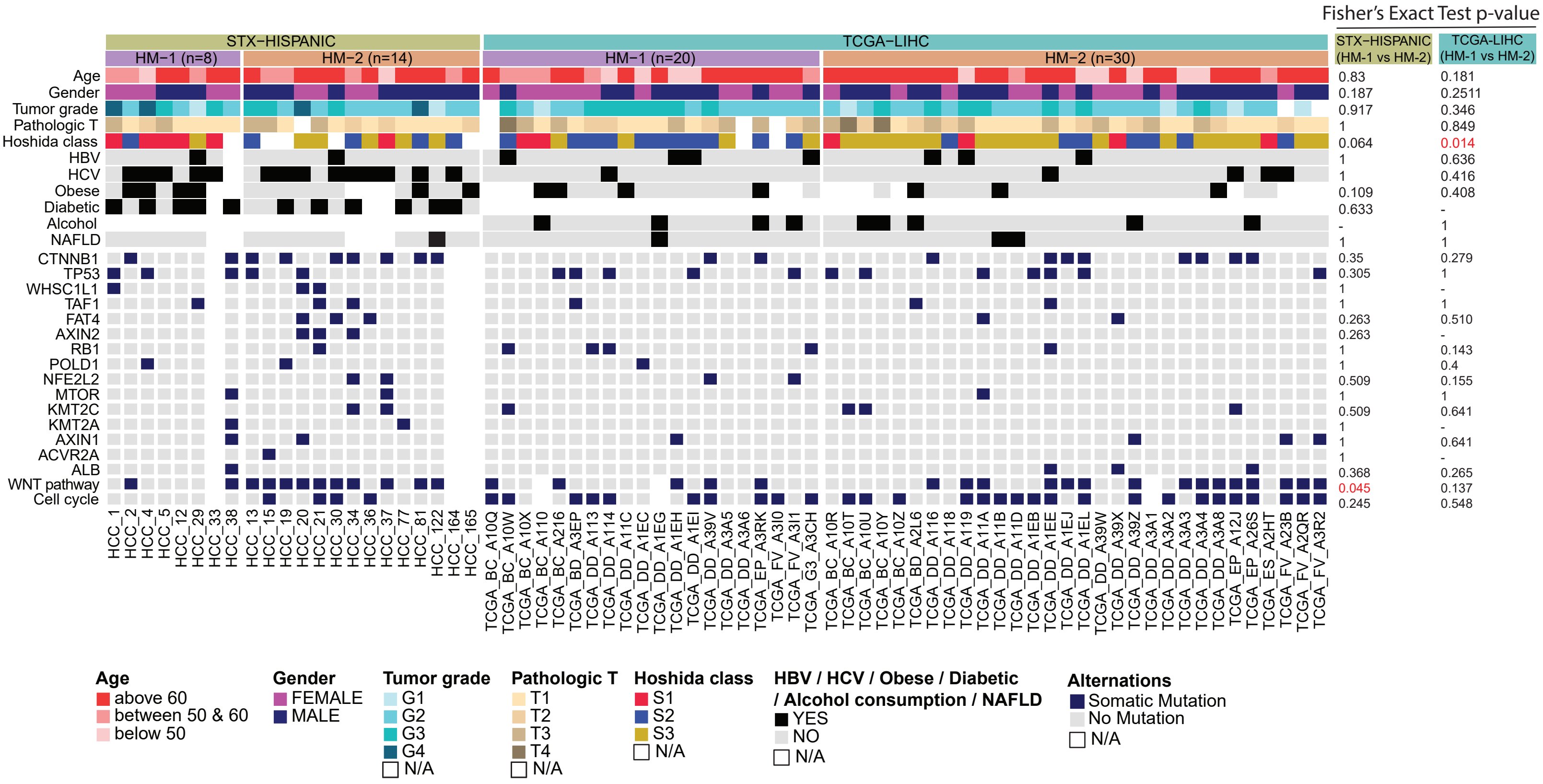


**Figure S4b:** Heatmap showing the selected clinical parameters, somatic mutations, and genomic alteration of oncogenic signaling pathways among the two clusters in STX-Hispanic HCC and TCGA-LIHC. We included 22 HCCs with consensus clustering information from transcriptome and proteome-based studies on our cohort. The p-value between HM-1 (n=8 for Hispanic HCC; n=20 for TCGA) and HM-2 (n=14 for Hispanic HCC; n=30 for TCGA) patients were calculated using Fisher's exact test.

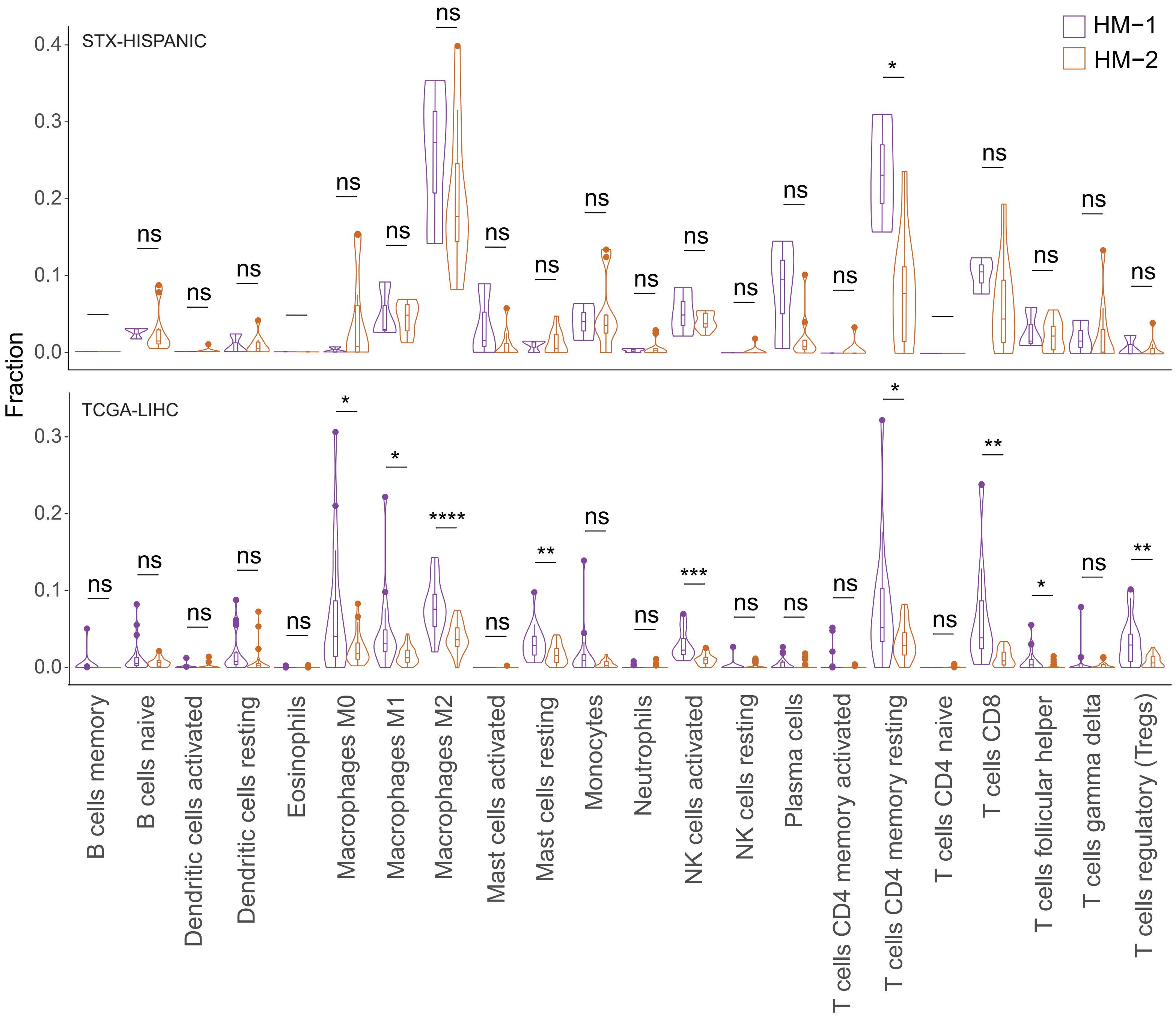


**Figure S5a:** Comparison of abundance of 22 immune cell types (LM22) between HM-1 and HM-2 tumors of the STX-Hispanic (top) and TCGA-LIHC (bottom) cohorts. The infiltration of the immune cell types was estimated by CIBERSORT absolute algorithm from RNA-Seq data. Welch's t-test was used to determine the significant difference between the means of the two abundances.

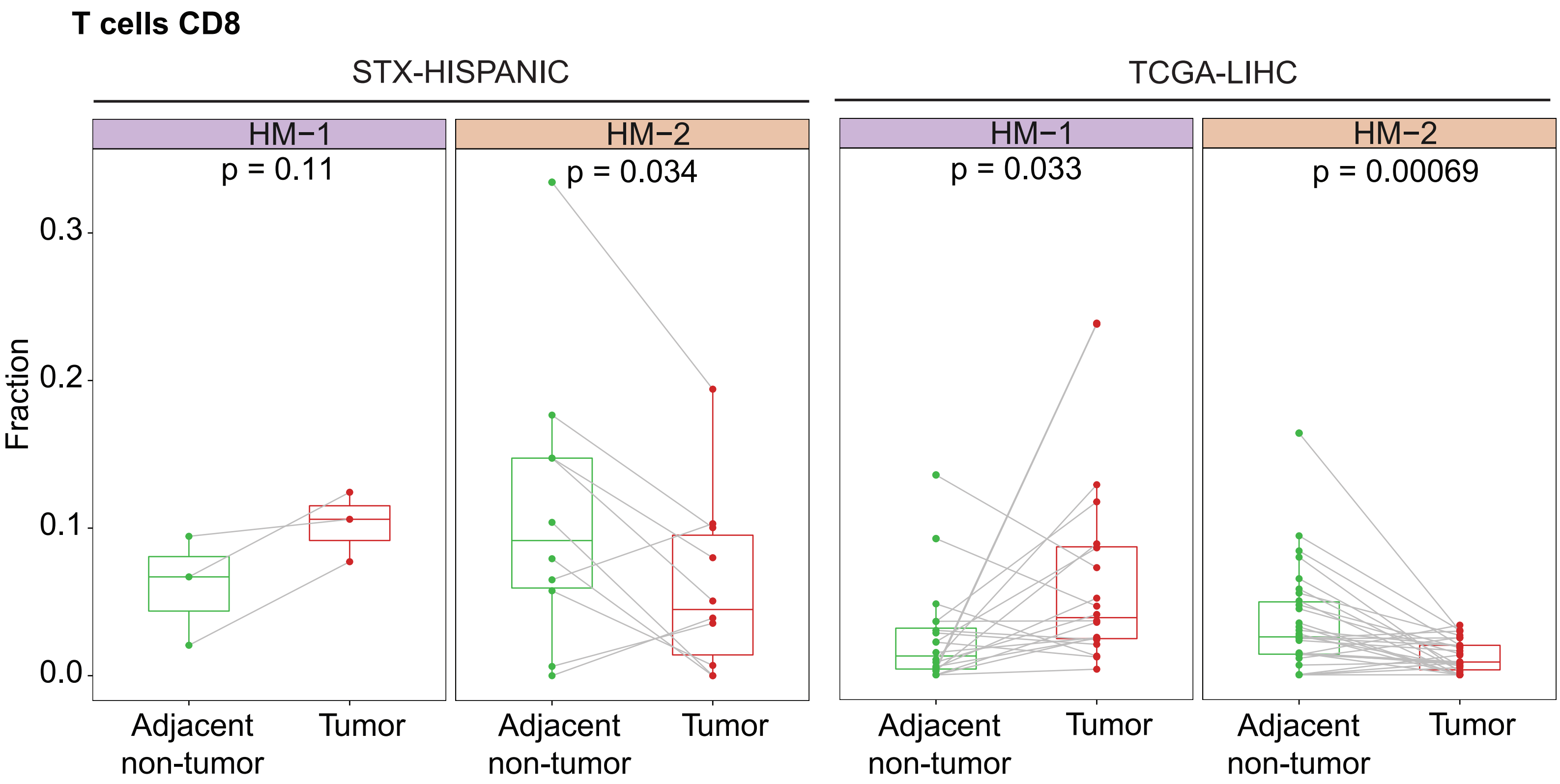


**Figure S5b:** Plots showing differential abundances of CD8+ T cells between tumor and paired non-tumor samples within each cluster. All p-values are calculated using paired sample t-tests.

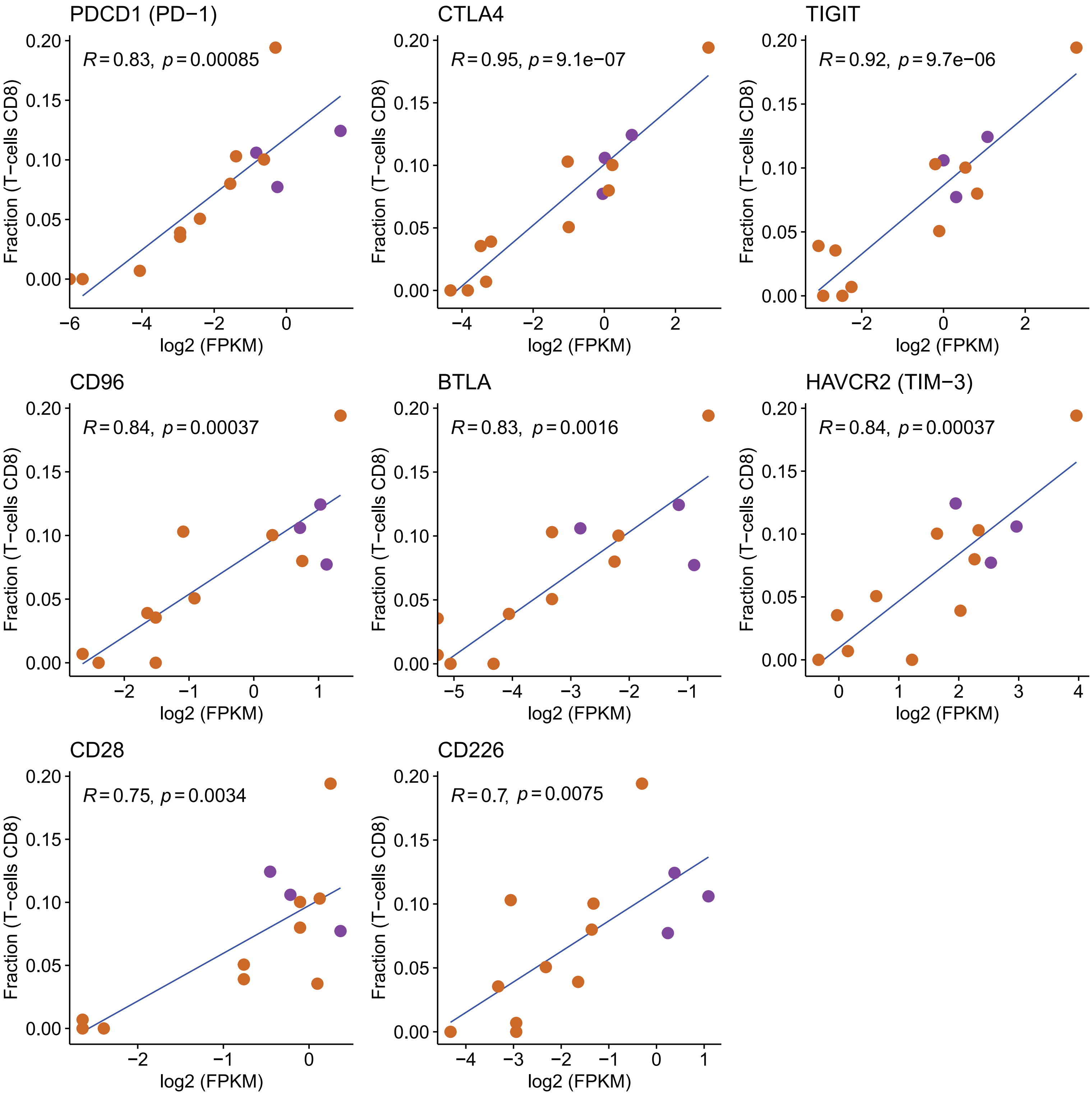


**Figure S5c:** Scatter plots showing the correlation between abundances of CD8+ T-cells (CIBERSORT absolute) and the expression (log2 FPKM) of eight cell surface marker genes in 13 HCC tumors of STX-Hispanic cohort. R values indicate Pearson's correlation coefficients.

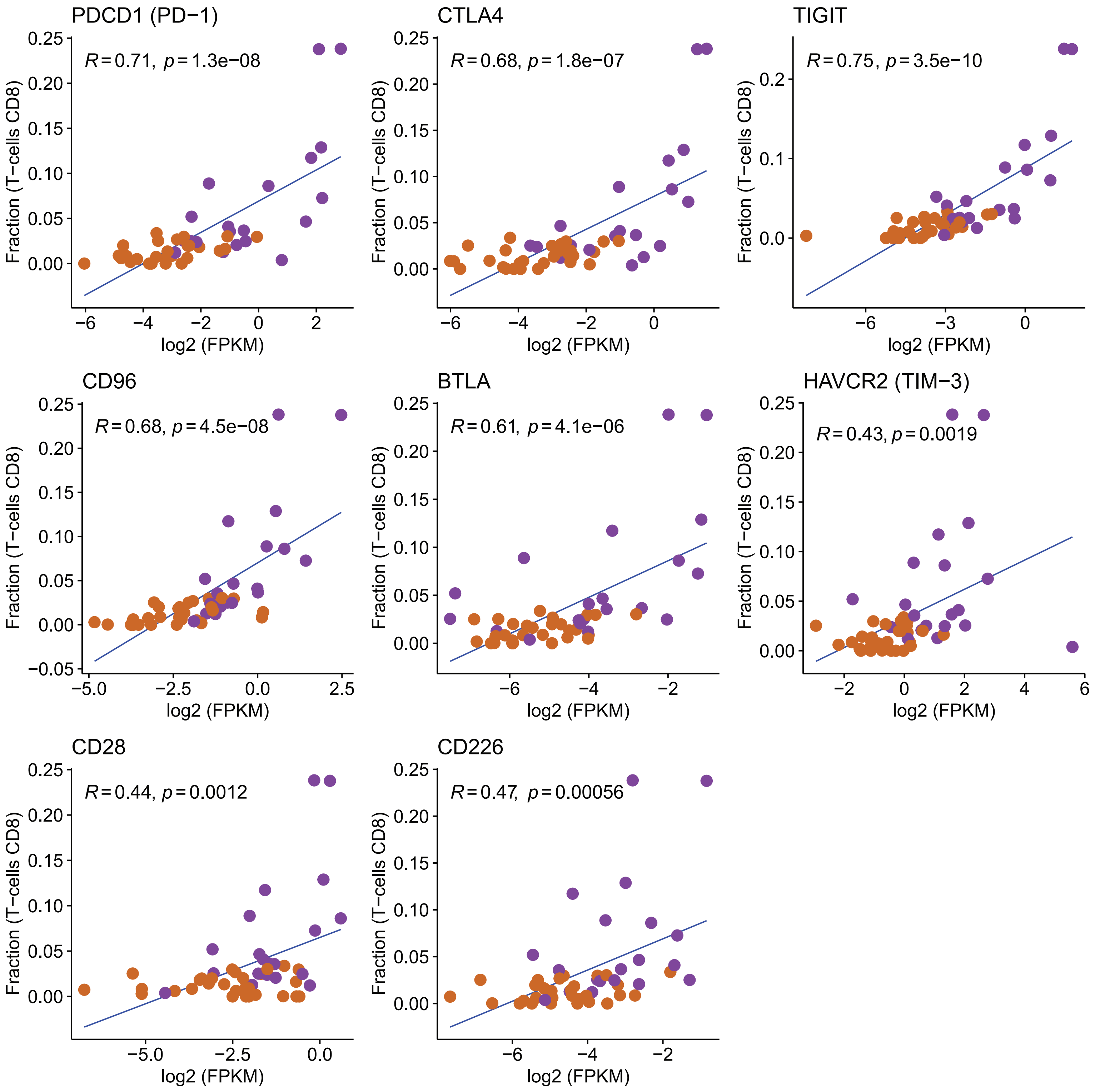


**Figure S5d:** Scatter plots showing the correlation between abundances of CD8+ T-cells (CIBER-SORT absolute) and the expression (log2 FPKM) of eight cell surface marker genes in 50 HCC tumors of the TCGA-LIHC cohort. R values indicate Pearson's correlation coefficients.

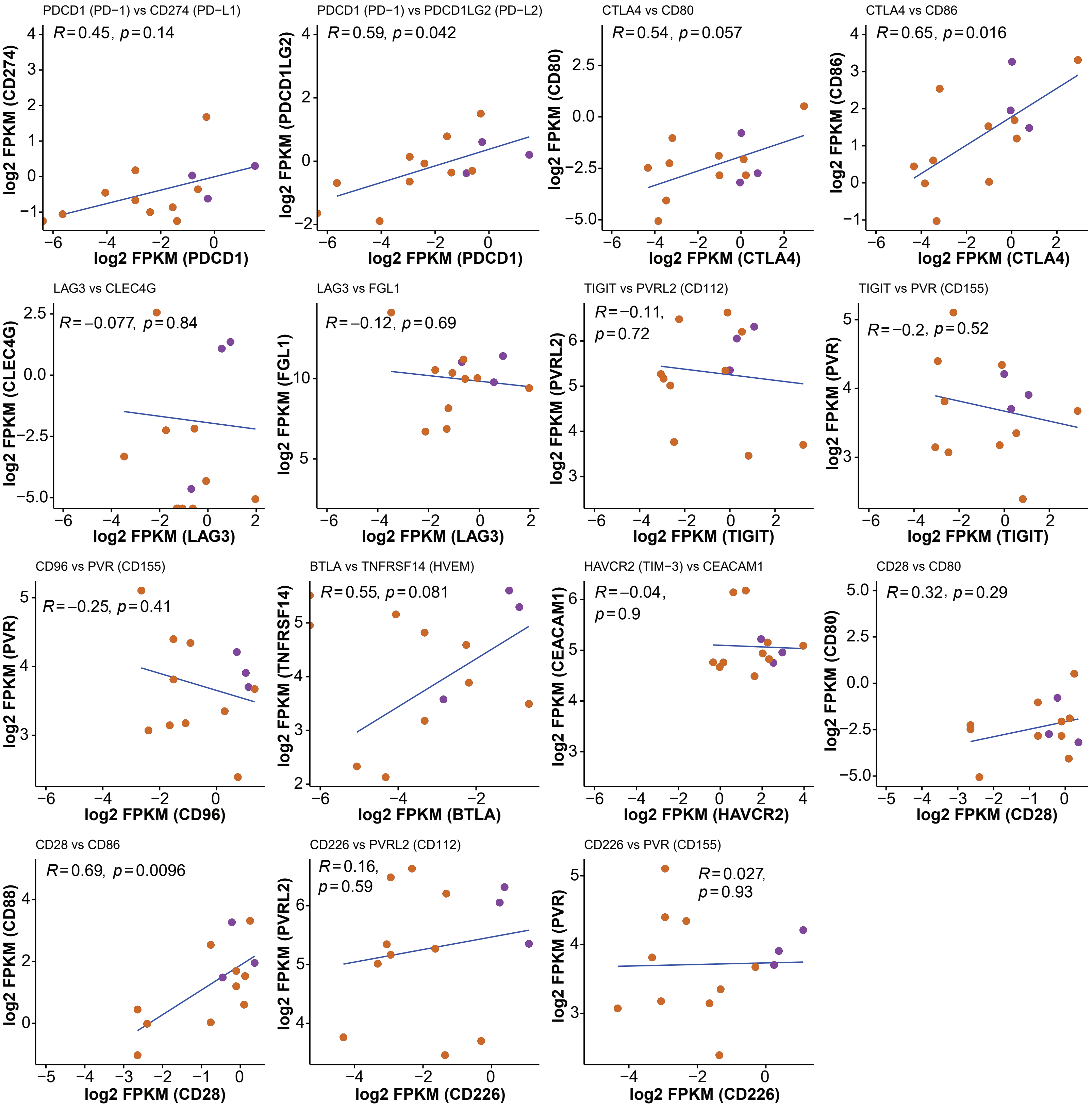


**Figure S5e:** Scatter plots for correlation between immune checkpoint molecules and their respective ligands in STX-Hispanic HCC tumor samples (n=13). The x- and y-axes indicate expressions (log2 FPKM) of immune checkpoint receptors and corresponding ligands, respectively. R values are indicative of Pearson's correlation coefficients.

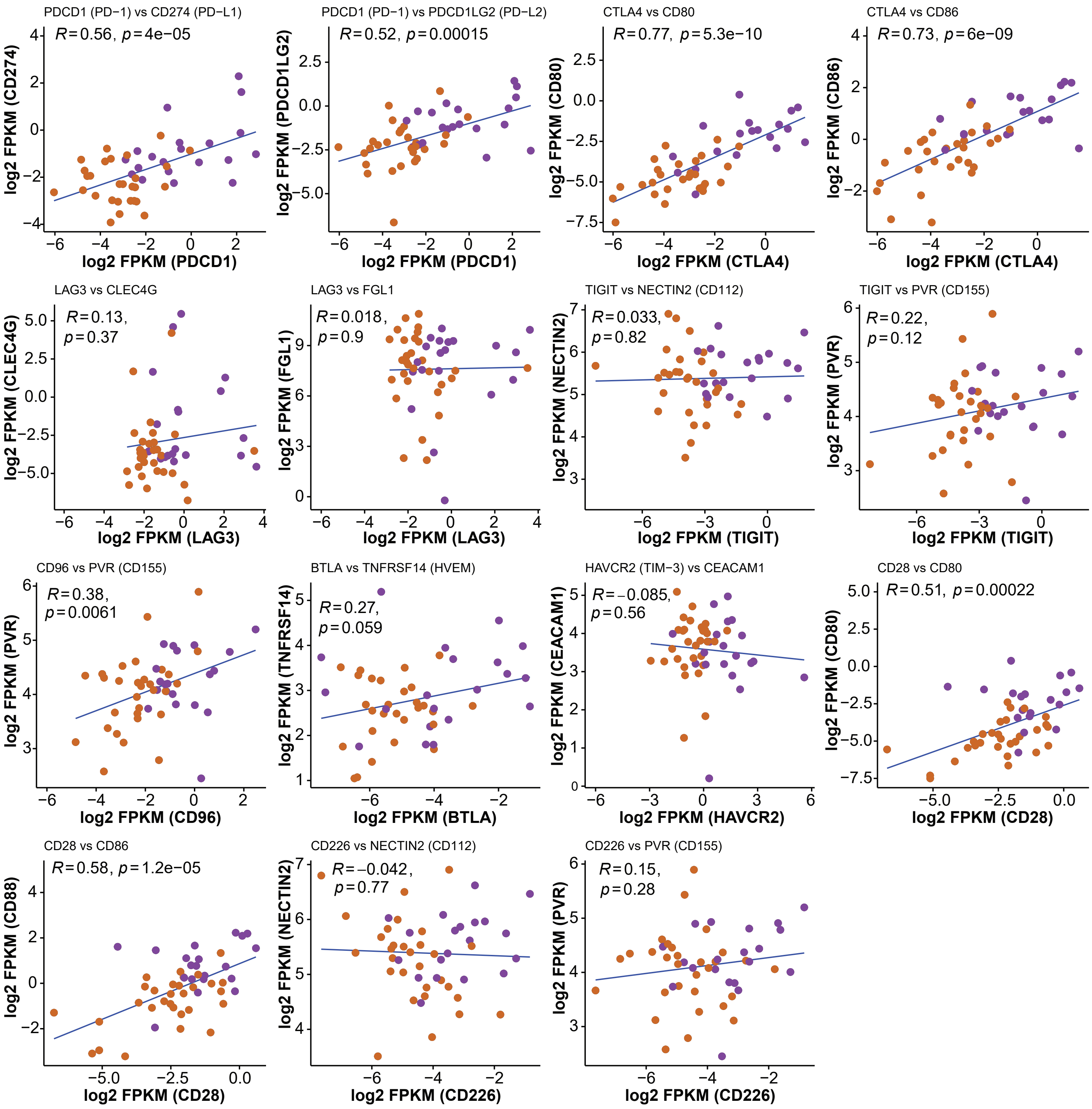


**Figure S5f:** Scatter plots for correlation between immune checkpoint molecules and their respective ligands in HCC tumor samples (n=50) from the TCGA-LIHC study. The x- and y-axes indicate expressions (log2 FPKM) of immune checkpoint receptors and corresponding ligands, respectively. R values are indicative of Pearson's correlation coefficients.

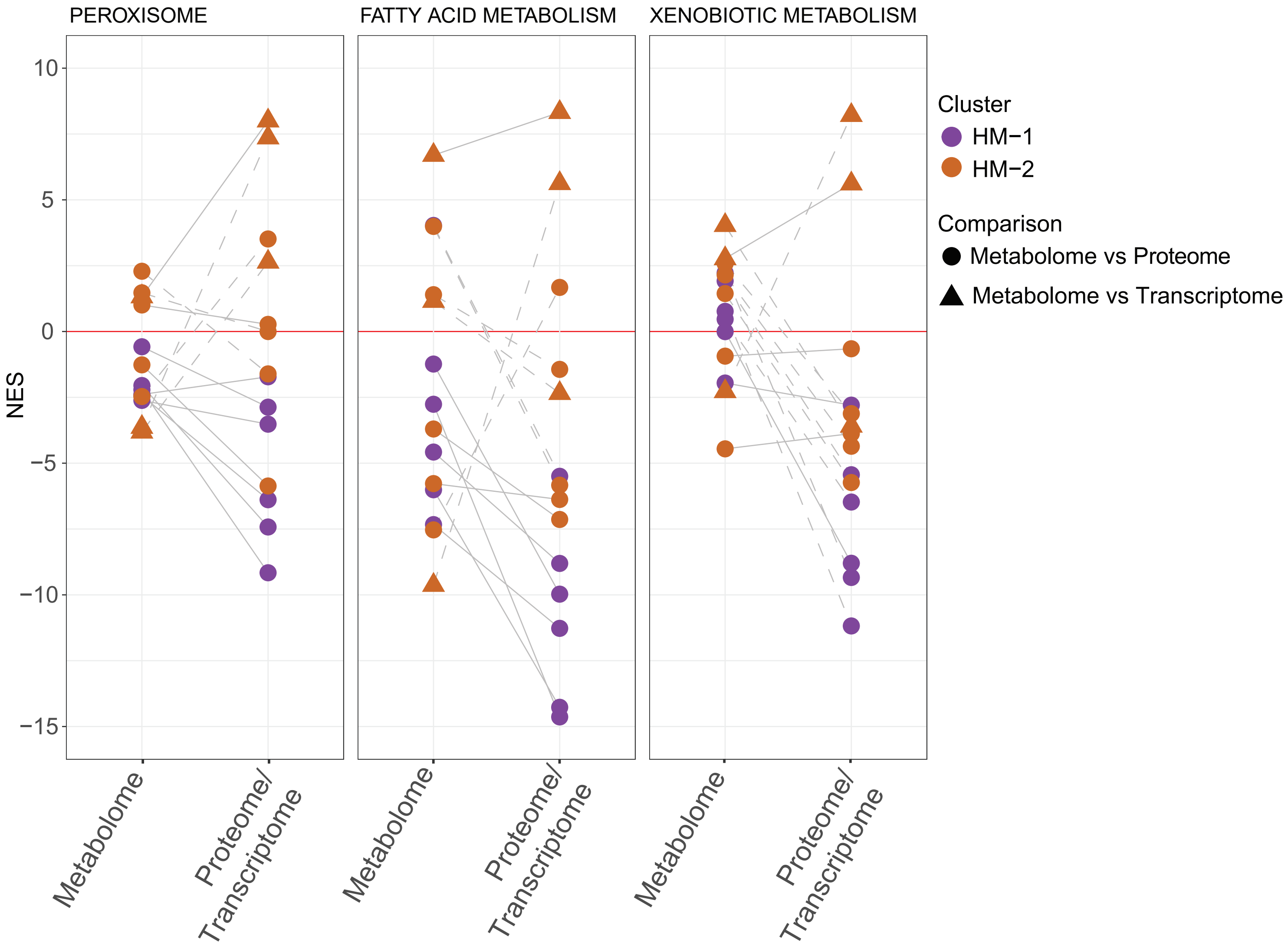


**Figure S6a:** Trends in enrichment scores (NES) of three proteome/transcriptome-based gene sets in metabolomic data of STX-Hispanic HCC (n=14). The solid lines indicate the concordance of enrichments and the dashed lines indicate difference in the directionality of enrichments between metabolomic and proteomic/transcriptomic data.

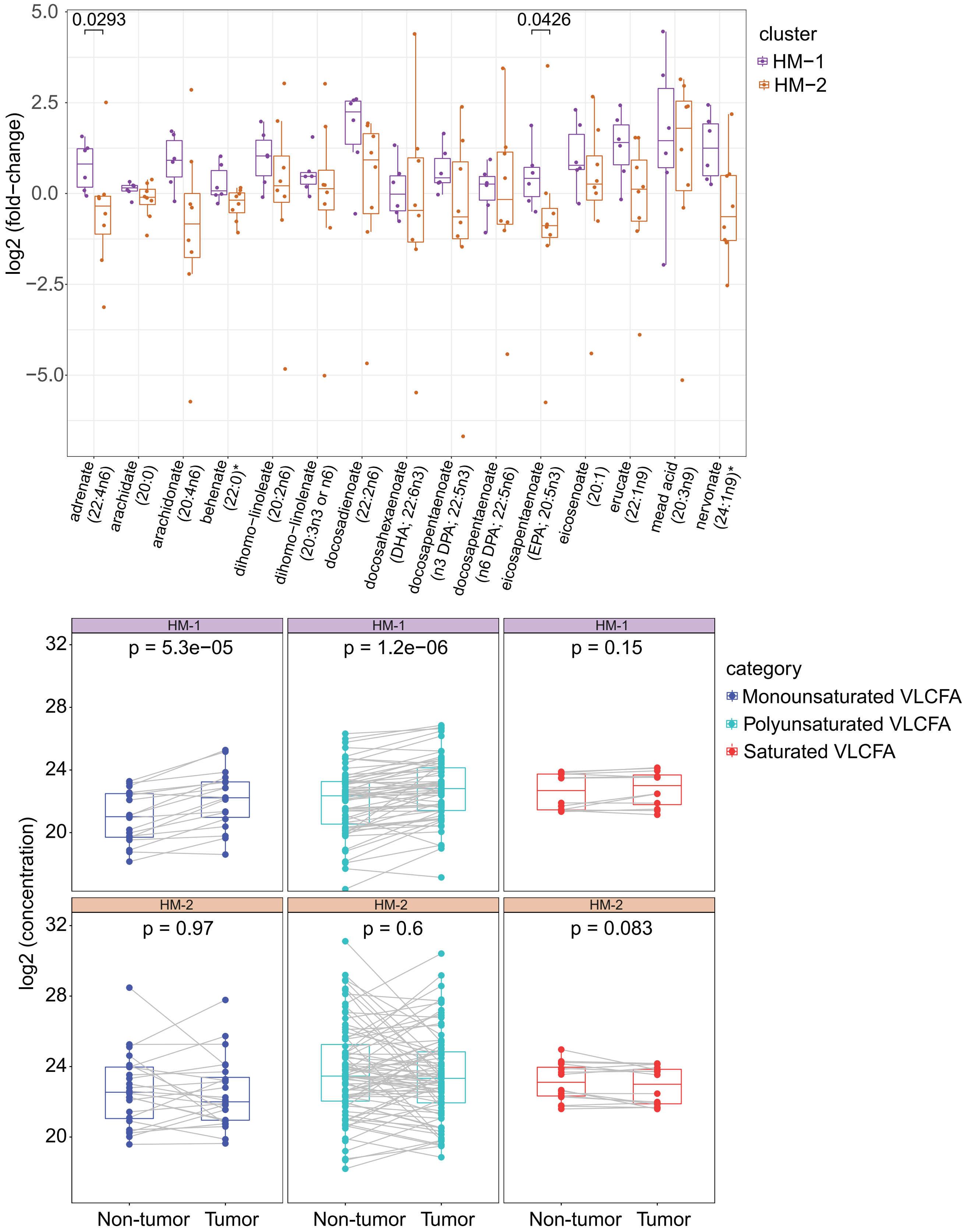


**Figure S6b:** Boxplot representation of log2 (fold-change) of selected very long-chain fatty acids (VLCFAs) in tumor compared to adjacent non-tumor samples of STX-Hispanic HCC (top panel). The VLCFAs have a chain length of ≥ 20 carbons. The bottom panel shows tumor-nontumor pairwise comparisons of all monounsaturated, polyunsaturated, and saturated VLCFAs in HM-1 (n=6) and HM-2 (n=8) patients. All p-values were calculated using Wilcoxon rank-sum (top panel) or signed-rank (bottom panel) tests.

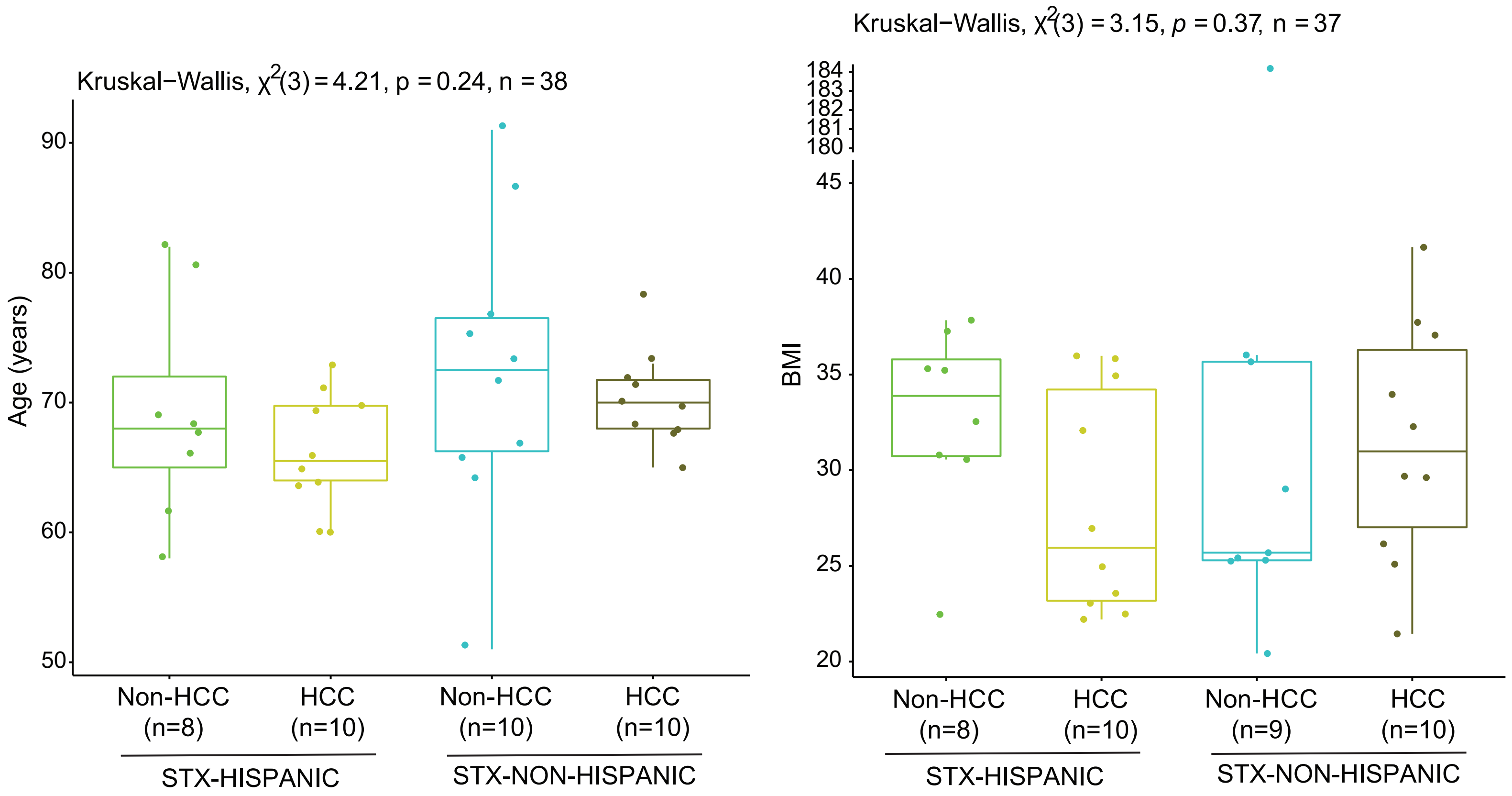


**Figure S7a:** Boxplots showing age distributions (in years) and BMI (body mass index) across the groups of HCC/non-HCC individuals recruited for our serum lipidomic study. We removed two outlier non-HCC samples before examining for any differences by the Kruskal−Wallis test.

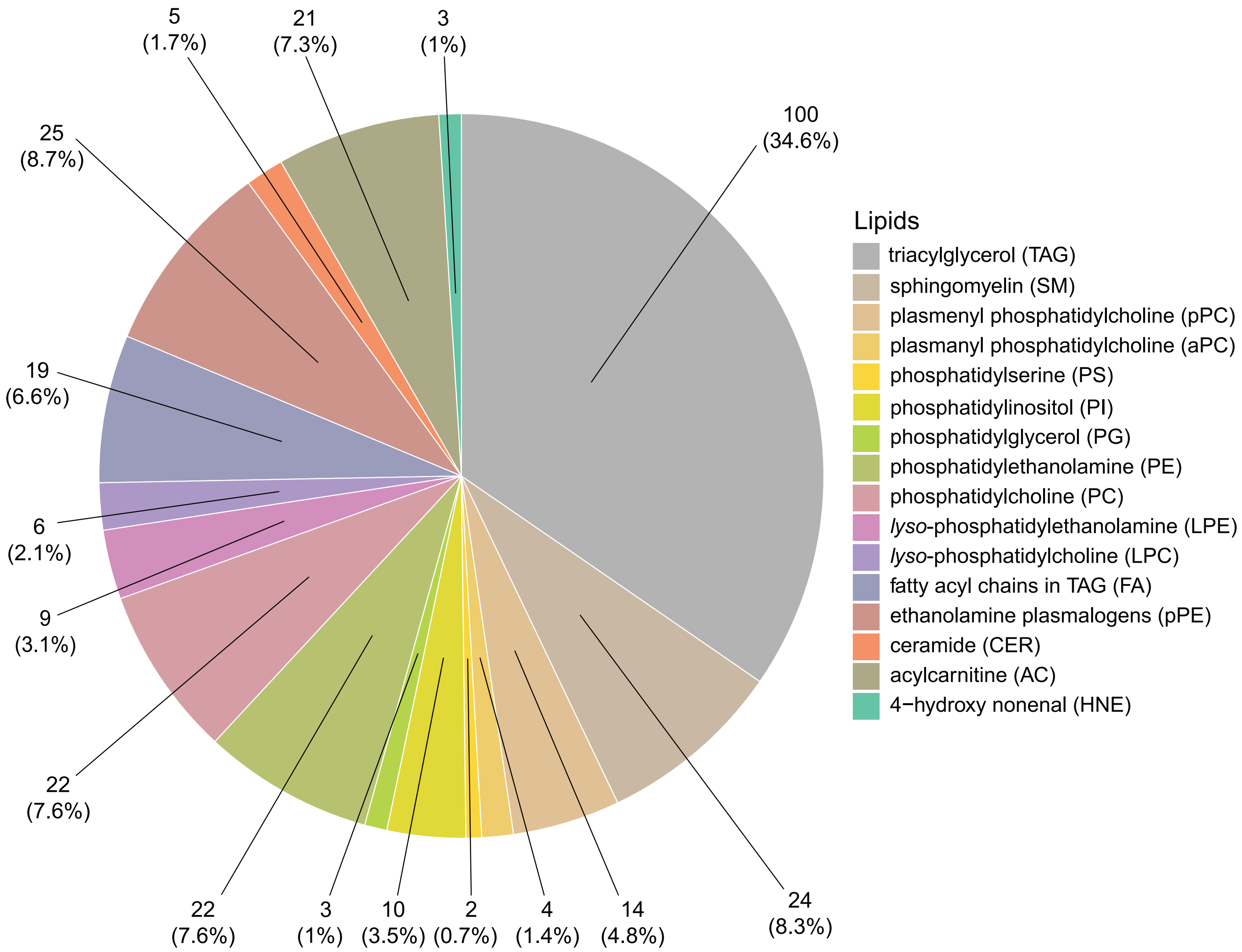


**Figure S7b:** Pie chart showing the different serum lipids included in our study. The proportion of each of the 16 profiled lipid types is indicated within the parenthesis.
